# Tracing the transmission of carbapenem-resistant *Enterobacterales* at the patient:ward environmental nexus

**DOI:** 10.1101/2024.07.03.24309291

**Authors:** Linzy Elton, Alan Williams, Shanom Ali, Jelena Heaphy, Vicky Pang, Liam Commins, Conor O’Brien, Özge Yetiş, Estelle Caine, Imogen Ward, Monika Muzslay, Samuel Yui, Kush Karia, Ellinor Shore, Sylvia Rofael, Damien Mack, Timothy D McHugh, Emmanuel Q Wey

## Abstract

**Introduction:** Colonisation and infection with Carbapenem-resistant Enterobacterales (CRE) in healthcare settings poses significant risks, especially for vulnerable patients. Genomic analysis can be used to trace transmission routes, supporting antimicrobial stewardship and informing infection control strategies. Here we used genomic analysis to track the movement and transmission of CREs within clinical and environmental samples.

**Methods:** 25 isolates were cultured from clinical patient samples or swabs, that tested positive for OXA-48-like variants using the NG-Test® CARBA-5 test and whole genome sequenced (WGS) using Oxford Nanopore Technologies (ONT). 158 swabs and 52 wastewater samples were collected from the ward environment. 60 isolates (matching clinical isolate genera; *Klebsiella, Enterobacter, Citrobacter and Escherichia*) were isolated from the environmental samples. Metagenomic sequencing was undertaken on 36 environmental wastewater and swab samples.

**Results:** 21/25 (84%) clinical isolates had >1 *bla_OXA_* gene and 19/25 (76%) harboured >1 *bla_NDM_* gene. Enterobacterales were most commonly isolated from environmental wastewater samples 27/52 (51.9%), then stick swabs 5/43 (11.6%) and sponge swabs 5/115 (4.3%). 11/60 (18%) environmental isolates harboured at >1 *bla_OXA_* gene and 1.9% (1/60) harboured *bla_NDM-1_*. *bla*_OXA_ genes were found in 2/36 (5.5%) metagenomic environmental samples.

**Discussion:** Potential for putative patient-patient and patient-ward transmission was shown. ONT sequencing can expedite clinical decisions whilst awaiting reference laboratory results, providing economic and patient care benefits. Metagenomic sampling needs optimization to improve sensitivity.

## 1 Introduction

Colonisation and subsequent healthcare associated infection (HCAI) with multi-drug resistant organisms (MDROs) is a concern for vulnerable patient groups, such as the elderly, or immunocompromised, within the hospital setting. Many of these HCAI incidents could be preventable with enhanced infection and control (IPC) measures [1]. The last line therapy for use against MDROs are the carbapenems and infection with carbapenem-resistant Enterobacterales (CRE) organisms are associated with high patient mortality rates [2]. CREs are difficult to treat as carbapenemase enzymes can hydrolyse almost all β-lactam antibiotics[3].

Oxacillinase-48-type carbapenemases (OXA-48) and New Delhi metallo-β-lactamase (NDM) are common CRE resistance mechanisms, which are now found globally, are highly mobile and no longer confined to the original bacterial species that they were characterised from [2], [4]. CRE resistance is most often transferred between bacterial isolates on mobile genetic elements, including plasmids, and a wide range of plasmid types have been seen in CRE organisms [4]. CREs are often associated with other genes that confer β-lactam resistances, such as *bla*_SHV_ and *bla*_CTX_, as they can be found on the same plasmids, complicating detection [5].

CREs have been found in both community settings and hospital environments around the globe [6]. Whilst colonised patients do not need antibiotic therapy, they still pose a transmission risk, and so both colonisation and infection must be considered when undertaking IPC. CREs have been isolated from high-touch surfaces, such as door handles, medical equipment trollies, as well as bed sheets and rails, and also from hospital wastewater, including sink U-bends [2], [7], [8], [9], [10]. The colonisation of hospital wastewater by CREs and other MDROs may be particularly problematic, as this water will pass out of the hospital and into the general wastewater system. Without adequate processing at wastewater treatment plants, and the risk of sewage being released into water systems, this may lead to the distribution of these MDROs and their AMR genes back into environmental and community settings [11], [12]. This is especially likely to be problematic in low- and middle-income settings, where the treatment of wastewater may not be adequate to remove these organisms [13].

Surveillance is crucial for the containment of pathogen and AMR outbreaks, especially in hospital settings with vulnerable patients and multiple-occupancy bed bays [14]. The current standard testing for CREs is culture and phenotypic antibiotic sensitivity testing (AST), and molecular methods such as PCR or rapid diagnostic tests (RDTs), such as lateral flow devices (including the NG-Test CARBA 5) [15].

As the COVID-19 pandemic showed with real-time tracking of variants, genomic analysis provides vital enhanced surveillance of transmission patterns [16]. Phenotypic and other molecular tests for resistance genes cannot identify the genetic relatedness of isolates, and thus cannot accurately track potential transmission. The use of whole genome sequencing (WGS) and metagenomics in the diagnosis and surveillance of MDROs can identify phylogeny, novel drug resistance mutations and inform design of targeted diagnostics [17].

*bla_OXA_*- and *bla_NDM_*-mediated CRE colonisations and infections were detected in several patients occupying single-bed rooms and multiple-occupancy bed bays on wards at a North London tertiary referral hospital between 2022-2023. Temporal and spatial associations between patients indicated the possibility of ongoing transmission events but routine phenotypic and molecular testing was unable to pinpoint transmission routes. In this study we aimed to use genomic analysis to build a picture of the movement and transmission of CRE species, AMR and plasmids within clinical pathogens and environmental species found in the ward environment.

## 2 Methods

### 2.1 Sample collection

Environmental sampling was requested by the hospital Trust and IPC lead for investigation as part of the extended standard of care. Further characterisation by molecular typing of the CREs that were part of the outbreak was included as an extension of the routine standard of diagnostic care pathway. Patient metadata was obtained through the electronic clinical infection database (elCID). AST profiles of the clinical bacterial pathogens isolated relevant to the CRE surveillance cases were undertaken following the EUCAST Clinical breakpoints (v12.0) for Gram Negative bacteria [18].

Twenty-eight clinical isolates were obtained from 20 patients with a positive NG-Test® CARBA-5 (NG Biotech Laboratories) immunochromatographic lateral flow test for *bla*_OXA_ and/or *bla*_NDM_ resistance on multiple wards between 7 February 2022 and 20 January 2023 [19]. Four were from infection sites and 24 were CRE screen samples. These CRE screens were rectal swabs, which were plated onto speciation agar, any suspected to be CREs were tested using the CARBA-5 lateral flow test, as per Health Services Laboratories ‘healthcare associated infection detection of carbapenemase producing organisms by culture’ standard operating procedure. Phenotypic pathogen species data collected from patient samples was obtained using the MALDI-TOF (Bruker). MALDI-TOF-MS of bacterial isolates was undertaken from pure isolates no older than 24hrs from culture. Isolates were spotted in duplicate and identifications with corresponding Log scores ≥2.0 “high-confidence to the species level” were considered only, and reported in the results. Patient spatio-temporal metadata was collected, for details please contact the authors.

For environmental sampling, a site visit to ward 7D was undertaken prior to collection, to evaluate ward layout and staff and patient routes of travels and sampling locations. Samples were taken from every multiple-occupancy bed bay and single-bed room, covering affected patient areas and non-affected areas. From each bay and room every sink and drain was sampled (in both bed and bathroom areas) and shower drains sampled. Non-clinical rooms were also covered, such as the shared use pantry, staff toilets, nurses station, storage rooms, sluice and workstations on wheels.

Two hundred µL of wastewater samples were collected into sterile sample containers pre-dosed with 1mL of a neutralising buffer comprising: 3% (w/v) Tween 80, 0.3% (w/v) Lecithin, 1.0% (w/v) Sodium thiosulfate, 1.5% (w/v) K_2_HPO_4_, KH_2_PO_4_ 0.05% (w/v), 1% (w/v) Poly-[sodium-4-styrenesulfonate], 0.1% (v/v) Triton® ×100 (Sigma-Aldrich, UK) and prepared in Phosphate-buffered saline (PBS) solution (Oxoid, UK) as previously described [20]. Samples were refrigerated (2-8°C) within 2 hours of collection and processed within 24 hours. Aliquots (0.5 mL, 0.1 mL) from the neat and serial 1/10 and 1/100 dilutions from the original wastewater samples were surface-plated onto selective agars: Colorex™ mSuperCARBA™ (EO Labs, UK), Brilliance CRE, *E. coli* Coliform, *Pseudomonas* CN (Oxoid, UK) and non-selective Columbia Blood Agar (Oxoid, UK). Plates were incubated aerobically at 37°C for 72 hours and inspected daily. A further 45 µL of the original samples were vacuum-filter concentrated as previously described [21] via 47mm diameter nitrocellulose membranes (0.45um pore size) and the membrane transferred to selective agars used above prior to incubation.

Cotton-tipped stick swabs (SS352, Appleton Woods) and sponge swabs (TS/15-B, Technical Service Consultants Ltd.), both pre-moistened with a neutraliser buffer were used to collect samples from difficult-to-access areas, as described above. Stick swab sampled were transferred to sterile universal tubes containing 9 mL of diluent buffer (saline), 1 mL of neutralising buffer and 3-5 glass beads. The swab contents were released by bead-washing (vortex mixing) for 30 seconds. Aliquots and serial dilutions of the resulting homogenised suspension was plated on selective and non-selective agars as above.

Suspect colonies were harvested for streak-purification onto non-selective agars and confirmed by MALDI-TOF mass spectrometry (MS) (Maldi-TOF Biotyper IVD system Bruker Daltronics). The remaining portion of the environmental samples were preserved in 500 µL 1x DNA/RNA Shield (Zymo Research Corporation).

### 2.2 DNA extraction and quantification

Clinical and environmental isolates were grown on Columbia horse blood plates (Oxoid Limited), then the DNA extracted using the DNeasy Blood & Tissue Miniprep Kit (Qiagen), following manufacturer’s instructions [22]. Metagenomics DNA from a subset of environmental samples, due to limited resources, was extracted directly from the environmental swab and water samples using the ZymoBIOMICS™ DNA Miniprep Kit (Zymo Research Corporation), following manufacturer’s instructions [23]. A sample of ZymoBIOMICS™ Microbial Community Standard (Zymo Research Corporation) was included, following manufacturer’s instructions [24]. DNA quality was assessed for concentration using the Qubit™ dsDNA BR Assay Kit (Thermo Fisher) and molecular weight and DNA integrity was confirmed using the Genomic DNA ScreenTape and reagents on the TapeStation 4150 (Agilent Technologies Inc.).

### 2.3 Library preparation

Clinical and environmental isolate DNA libraries were prepared using the Rapid Barcoding Kit 96 (SQK-RBK110.96) with a DNA input of between 50-200 ng, following manufacturer’s instructions [25]. For the environmental swab samples, metagenomic DNA libraries were prepared using the ONT Rapid PCR Barcoding Kit (SQK-RPB004) with a DNA input of 1-5 ng and following the manufacturers’ instructions [26]. ZymoBIOMICS™ Microbial Community DNA Standards (Zymo Research Corporation) were included, following manufacturer’s instructions [27].

### 2.4 Sequencing and basecalling

Up to 24 barcoded clinical and environmental isolate samples, or 12 barcoded environmental metagenomic samples were run together on a flow cell version R9.4.1 (Oxford Nanopore Technologies) using a MinION device for 72 hours, using the default parameters on the MinKNOW software (v23.04.6). Basecalling was performed either by the MinKNOW software alongside sequencing or using the Guppy basecalling software (v6.5.7) [41], using the flip-flop high accuracy algorithm, with a minimum Q score of 8 and minimum depth of 40x.

### 2.5 Data analysis

Fastq files were quality checked (QC) using FastQC (v0.21.1) and MultiQC (v1.15) and those with a read depth of >40x were included in further analysis [28], [29]. 25 clinical isolates, 60 environmental isolates, 36 metagenomics environmental samples were included in this analysis. Barcodes were trimmed from the reads using Guppy. All clinical and environmental isolate, and environmental metagenomic samples were analysed for the presence of AMR genes using KmerResistance 2.2 [30], [31] and for species using KmerFinder 3.2 (v3.0.2) [30], [32], [33]. Plasmids were identified using PlasmidFinder 2.1 (v2.0.1) [30], [34].

Clinical and environmental isolate fastq files were aligned to the reference genome for their species (*C. freundii*: GCF_003812345.1, *C. portucalensis*: GCA_023374935.1, *C. youngae*: GCF_015139575.1, *E. cloacae*: GCF_905331265.2, *E. bugandensis*: GCF_020042625.1, *E. hormaechei*: GCF_024218835.1, *E. coli*: GCF_000005845.2, *K. pneumoniae*: GCF_000240185.1, *K. michigenensis*: GCF_015139575.1) using MiniMap2 (v2.26) [35], then sorted and indexed using Samtools (v1.17) [36]. Alignments were visualised using Artemis (v18.1.0) [37]. Depth and coverage were calculated using Samtools. Consensus fasta files were created using Samtools and then dendograms for each species (with three or more isolates) were created using Parsnp (utilising maximal unique matches) (v1.7.4) [38], [39], [40], [41] and visualised using iTOL (v6) [42].

Speciation for the results and discussion were as per the genomic speciation. Sequence data were deposited under BioProject PRJEB76684 on the European Nucleotide Archive and outlined in supplementary materials S1, S2 and S3.

## 3 Results

### 3.1 MALDI-TOF vs WGS for isolate speciation

All 25 clinical and 48 environmental isolates were subjected to both MALDI and WGS analysis. Non-concordant *C. freundii* by MALDI-TOF were speciated as *C. portucalensis* (one clinical isolate and one environmental isolate) or *C. youngae* (three environmental isolates) in WGS. One MALDI-TOF call of *C. braakii*/*freundii* was called as *C. youngae* using WGS. One environmental isolate that MALDI-TOF identified as *C. freundii* was speciated as *P. mirabilis* (possibly a mixed culture). Non-matching *E. cloacae* were speciated as *E. asburiae* (one environmental isolate) and *E. hormaechei* (nine, including all five of the clinical isolates and four environmental isolates). All MALDI-TOF and WGS speciation for *E. coli* and *K. pneumoniae* was concordant. All the MALDI-TOF-called *K. oxytoca* were speciated as either *K. michigenensis* (4/5 isolates) or *K. grimontii* (1/5 isolates) using WGS. See Table 1 and supplementary materials S2.

**Table 1.**
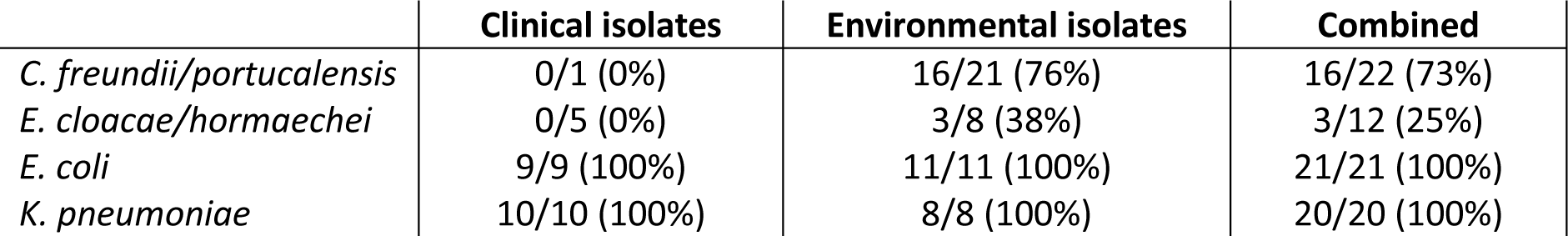
Concordance of MALDI-TOF speciation vs WGS for both clinical and environmental isolates. All non-concordant samples were either of the *C. freundii* or *E. cloacae* complex. Note that 48/60 environmental isolates are included in this table, the rest were different species.

|  | Clinical isolates | Environmental isolates | Combined |
| --- | --- | --- | --- |
| *C. freundii/portucalensis* | 0/1 (0%) | 16/21 (76%) | 16/22 (73%) |
| *E. cloacae/hormaechei* | 0/5 (0%) | 3/8 (38%) | 3/12 (25%) |
| *E. coli* | 9/9 (100%) | 11/11 (100%) | 21/21 (100%) |
| *K. pneumoniae* | 10/10 (100%) | 8/8 (100%) | 20/20 (100%) |

### 3.2 Environmental sampling

210 environmental samples were collected, including 43 stick swabs, 115 sponge swabs and 52 wastewater samples. In total, 195 bacteria were isolated from 36 (16.9%) environmental samples, 76 isolates were Enterobacterales: 6 from stick swabs, 5 from sponge swabs and 65 from non-potable water samples. There was a significant difference in the proportion of samples from which Enterobacterales species were isolated from when the Chi-square test was applied *p*=<0.0001. Enterobacterales were isolated from 51.9% of water samples, compared with 11.6% of stick swabs and 4.3% of sponge swabs (see Table 2).

**Table 2.** Environmental sample types collected, and the number of each sample type from which Enterobacterales were isolated. Significance was calculated using Chi-Squared test.

| Environmental sample type | Total number of samples | Number from which Enterobacterales were isolated | Number from which no Enterobacterales were isolated | Significance (Chi-squared) |
| --- | --- | --- | --- | --- |
| Stick swab | 43 | 5 (11.6%) | 38 (88.3%) | $p < 0.0001$ |
| Sponge swab | 115 | 5 (4.3%) | 110 (95.6%) |  |
| Water | 52 | 27 (51.9%) | 25 (48.1%) |  |
| Total samples | 210 | 37 (17.6%) | 173 (82.3%) |  |

Enterobacterales were isolated from 36 different environmental samples (mean = 2.2 isolates per site, SD = 1.4). See Table 3. Table 4Table *5* details the sample sites from which each species was isolated, Table 5 describes the ward areas in which they were isolated. For full data see supplementary materials S3.

**Table 3.**
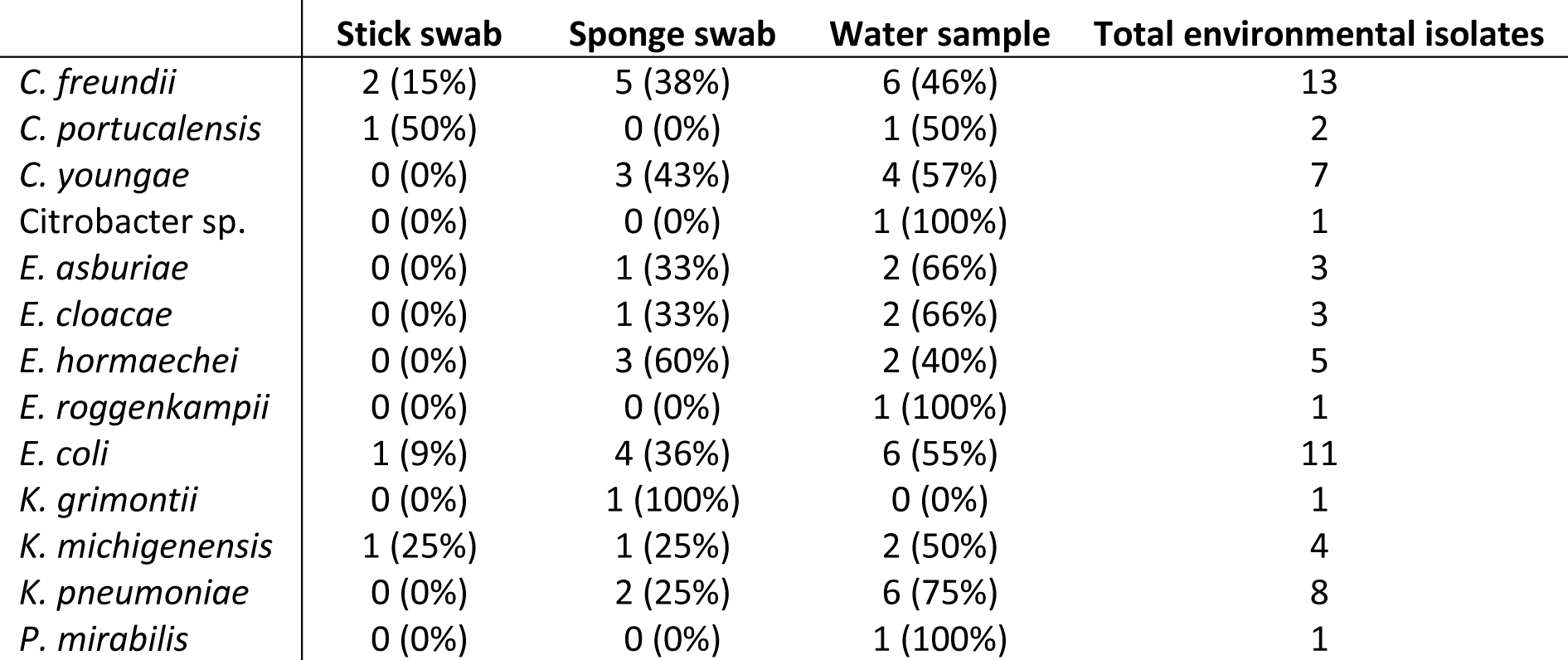
number of each species isolated from each different type of environmental sample.

|  | Stick swab | Sponge swab | Water sample | Total environmental isolates |
| --- | --- | --- | --- | --- |
| *C. freundii* | 2 (15%) | 5 (38%) | 6 (46%) | 13 |
| *C. portucalensis* | 1 (50%) | 0 (0%) | 1 (50%) | 2 |
| *C. youngae* | 0 (0%) | 3 (43%) | 4 (57%) | 7 |
| *Citrobacter* sp. | 0 (0%) | 0 (0%) | 1 (100%) | 1 |
| *E. asburiae* | 0 (0%) | 1 (33%) | 2 (66%) | 3 |
| *E. cloacae* | 0 (0%) | 1 (33%) | 2 (66%) | 3 |
| *E. hormaechei* | 0 (0%) | 3 (60%) | 2 (40%) | 5 |
| *E. roggenkampii* | 0 (0%) | 0 (0%) | 1 (100%) | 1 |
| *E. coli* | 1 (9%) | 4 (36%) | 6 (55%) | 11 |
| *K. grimontii* | 0 (0%) | 1 (100%) | 0 (0%) | 1 |
| *K. michigenensis* | 1 (25%) | 1 (25%) | 2 (50%) | 4 |
| *K. pneumoniae* | 0 (0%) | 2 (25%) | 6 (75%) | 8 |
| *P. mirabilis* | 0 (0%) | 0 (0%) | 1 (100%) | 1 |

**Table 4.** Environmental isolates present divided by sample site and type depicting an environmental reservoir density by location. HWB = hand wash basin, DWT = drain waste trap, WoW = workstation on wheels.

| Sample site and type | *C. freundii* | *C. portucalensis* | *C. youngae* | *Citrobacter* sp. | *E. asburiae* | *E. cloacae* | *E. hormaechei* | *E. roggkampii* | *E. coli* | *K. grimontii* | *K. michigenensis* | *K. pneumoniae* | *P. mirabilis* | Total species found per site |
| --- | --- | --- | --- | --- | --- | --- | --- | --- | --- | --- | --- | --- | --- | --- |
| Bathroom HWB drain | 1 (50%) | 0 (0%) | 0 (0%) | 0 (0%) | 0 (0%) | 0 (0%) | 0 (0%) | 0 (0%) | 1 (50%) | 0 (0%) | 0 (0%) | 0 (0%) | 0 (0%) | 2 |
| Bathroom HWB DWT | 1 (17%) | 0 (0%) | 2 (33%) | 1 (17%) | 1 (17%) | 1 (0%) | 0 (0%) | 0 (0%) | 0 (0%) | 0 (0%) | 0 (0%) | 1 (17%) | 0 (0%) | 7 |
| Bathroom toilet bowl | 1 (25%) | 1 (25%) | 0 (0%) | 0 (0%) | 0 (0%) | 0 (0%) | 0 (0%) | 0 (0%) | 1 (25%) | 0 (0%) | 1 (25%) | 0 (0%) | 0 (0%) | 4 |
| Bathroom toilet floor | 1 (100%) | 0 (0%) | 0 (0%) | 0 (0%) | 0 (0%) | 0 (0%) | 0 (0%) | 0 (0%) | 0 (0%) | 0 (0%) | 0 (0%) | 0 (0%) | 0 (0%) | 1 |
| Medication Cupboard exterior surface | 0 (0%) | 0 (0%) | 0 (0%) | 0 (0%) | 0 (0%) | 0 (0%) | 0 (0%) | 0 (0%) | 0 (0%) | 0 (0%) | 0 (0%) | 0 (0%) | 0 (0%) | 0 |
| Domestic Waste skip lid | 0 (0%) | 0 (0%) | 0 (0%) | 0 (0%) | 0 (0%) | 0 (0%) | 1 (100%) | 0 (0%) | 0 (0%) | 0 (0%) | 0 (0%) | 0 (0%) | 0 (0%) | 1 |
| Drinks Cooler mixed Pre-Flush | 0 (0%) | 0 (0%) | 0 (0%) | 0 (0%) | 0 (0%) | 0 (0%) | 1 (100%) | 0 (0%) | 0 (0%) | 0 (0%) | 0 (0%) | 0 (0%) | 0 (0%) | 1 |
| Bay HWB DWT | 9 (26%) | 0 (0%) | 5 (14%) | 0 (0%) | 1 (3%) | 3 (9%) | 0 (0%) | 1 (3%) | 6 (17%) | 0 (0%) | 2 (6%) | 7 (20%) | 1 (3%) | 35 |
| Office HWB DWT | 0 (0%) | 1 (50%) | 0 (0%) | 0 (0%) | 0 (0%) | 0 (0%) | 0 (0%) | 0 (0%) | 1 (50%) | 0 (0%) | 0 (0%) | 0 (0%) | 0 (0%) | 2 |
| Sink drain U bend exterior | 0 (0%) | 0 (0%) | 0 (0%) | 0 (0%) | 0 (0%) | 0 (0%) | 1 (100%) | 0 (0%) | 0 (0%) | 0 (0%) | 0 (0%) | 0 (0%) | 0 (0%) | 1 |
| Sink DWT | 0 (0%) | 0 (0%) | 0 (0%) | 0 (0%) | 1 (25%) | 0 (0%) | 0 (0%) | 0 (0%) | 1 (25%) | 1 (25%) | 1 (25%) | 0 (0%) | 0 (0%) | 4 |
| Staff toilet HWB DWT | 0 (0%) | 0 (0%) | 0 (0%) | 0 (0%) | 0 (0%) | 0 (0%) | 0 (0%) | 0 (0%) | 1 (100%) | 0 (0%) | 0 (0%) | 0 (0%) | 0 (0%) | 1 |
| WoW Keyboard and Mouse | 0 (0%) | 0 (0%) | 0 (0%) | 0 (0%) | 0 (0%) | 0 (0%) | 1 (100%) | 0 (0%) | 0 (0%) | 0 (0%) | 0 (0%) | 0 (0%) | 0 (0%) | 1 |
| Total isolates of each species present | 13 | 2 | 7 | 1 | 3 | 3 | 4 | 1 | 11 | 1 | 4 | 8 | 1 |  |

**Table 5.** Environmental isolates present divided by room indicating the environmental reservoir densities by room-type. Percentages describe the proportion each species makes up per location.

| Sample site and type | *C. freundii* | *C. portucalensis* | *C. youngae* | *Citrobacter* sp. | *E. asburiae* | *E. cloacae* | *E. hormaechei* | *E. roggenkampii* | *E. coli* | *K. grimontii* | *K. michigenensis* | *K. pneumoniae* | *P. mirabilis* | Total species found per site |
| --- | --- | --- | --- | --- | --- | --- | --- | --- | --- | --- | --- | --- | --- | --- |
| Bay (Beds 1-4) | 3 (60%) | 0 (0%) | 0 (0%) | 0 (0%) | 0 (0%) | 0 (0%) | 0 (0%) | 0 (0%) | 1 (20%) | 0 (0%) | 0 (0%) | 1 (20%) | 1 (20%) | 5 |
| Bay (Beds 15-18) | 1 (8%) | 0 (0%) | 2 (17%) | 1 (8%) | 0 (0%) | 3 (25%) | 0 (0%) | 0 (0%) | 2 (17%) | 0 (0%) | 2 (17%) | 1 (8%) | 0 (0%) | 12 |
| Bay (Beds 21-24) | 0 (0%) | 1 (50%) | 0 (0%) | 0 (0%) | 0 (0%) | 0 (0%) | 0 (0%) | 0 (0%) | 0 (0%) | 0 (0%) | 1 (50%) | 0 (0%) | 0 (0%) | 2 |
| Bay (Beds 29-32) | 4 (67%) | 0 (0%) | 1 (17%) | 0 (0%) | 0 (0%) | 0 (0%) | 0 (0%) | 0 (0%) | 0 (0%) | 0 (0%) | 0 (0%) | 1 (17%) | 0 (0%) | 6 |
| Bay (Beds 5-8) | 1 (25%) | 0 (0%) | 0 (0%) | 0 (0%) | 0 (0%) | 0 (0%) | 0 (0%) | 0 (0%) | 0 (0%) | 0 (0%) | 0 (0%) | 3 (75%) | 0 (0%) | 4 |
| Bed 9 | 1 (25%) | 0 (0%) | 0 (0%) | 0 (0%) | 0 (0%) | 0 (0%) | 0 (0%) | 1 (25%) | 1 (25%) | 0 (0%) | 0 (0%) | 1 (25%) | 0 (0%) | 4 |
| Bed 10 | 2 (50%) | 0 (0%) | 0 (0%) | 0 (0%) | 1 (25%) | 0 (0%) | 1 (25%) | 0 (0%) | 0 (0%) | 0 (0%) | 0 (0%) | 0 (0%) | 0 (0%) | 4 |
| Bed 19 | 0 (0%) | 0 (0%) | 2 (40%) | 0 (0%) | 0 (0%) | 0 (0%) | 0 (0%) | 0 (0%) | 2 (40%) | 0 (0%) | 0 (0%) | 1 (20%) | 0 (0%) | 5 |
| Bed 20 | 1 (25%) | 0 (0%) | 2 (50%) | 0 (0%) | 1 (25%) | 0 (0%) | 0 (0%) | 0 (0%) | 0 (0%) | 0 (0%) | 0 (0%) | 0 (0%) | 0 (0%) | 4 |
| Drug prep room | 0 (0%) | 0 (0%) | 0 (0%) | 0 (0%) | 0 (0%) | 0 (0%) | 1 (100%) | 0 (0%) | 0 (0%) | 0 (0%) | 0 (0%) | 0 (0%) | 0 (0%) | 1 |
| Office | 0 (0%) | 1 (25%) | 0 (0%) | 0 (0%) | 0 (0%) | 0 (0%) | 0 (0%) | 0 (0%) | 3 (75%) | 0 (0%) | 0 (0%) | 0 (0%) | 0 (0%) | 4 |
| Pantry (Staff & Patient use) | 0 (0%) | 0 (0%) | 0 (0%) | 0 (0%) | 1 (20%) | 0 (0%) | 1 (20%) | 0 (0%) | 1 (20%) | 1 (20%) | 1 (20%) | 0 (0%) | 0 (0%) | 5 |
| Sluice | 0 (0%) | 0 (0%) | 0 (0%) | 0 (0%) | 0 (0%) | 0 (0%) | 1 (100%) | 0 (0%) | 0 (0%) | 0 (0%) | 0 (0%) | 0 (0%) | 0 (0%) | 1 |
| Staff toilet | 0 (0%) | 0 (0%) | 0 (0%) | 0 (0%) | 0 (0%) | 0 (0%) | 0 (0%) | 0 (0%) | 1 (100%) | 0 (0%) | 0 (0%) | 0 (0%) | 0 (0%) | 1 |
| Workstation on wheels | 0 (0%) | 0 (0%) | 0 (0%) | 0 (0%) | 0 (0%) | 0 (0%) | 1 (100%) | 0 (0%) | 0 (0%) | 0 (0%) | 0 (0%) | 0 (0%) | 0 (0%) | 1 |
| Total isolates of each species found | 13 | 2 | 7 | 1 | 3 | 3 | 5 | 1 | 11 | 1 | 4 | 8 | 1 |  |

Table 6 details the environmental sample swab types, total number of bacterial species, and the total number of reads obtained from each of the 36 metagenomic sequencing sample sites that Enterobacterales were also present. There was no significant difference in the number of sequencing reads obtained, when comparing the swab type, or when comparing the swab location type. When comparing rooms, there was no significant differences, apart from Bay (beds 1-4) and bed 20 (*p*=0.0321). All tests were undertaken using One Way ANOVA and Tukey’s multiple comparison test. Speciation was based on genomic data, so only those that passed QC are included in the analysis.

**Table 6.** Environmental swab metagenomic samples, from which Enterobacterales were isolated, showing which type of sample was taken (sponge swab, stick swab or wastewater sample), number of bacterial species identified (including Gram negatives and Gram positives), and number of reads obtained from metagenomic sequencing. HWB = hand wash basin, DWT = drain waste trap.

| Environmental sample number | Room | Swab location | Total species found | Swab type | Total reads |
| --- | --- | --- | --- | --- | --- |
| 15 | Sluice | Domestic Waste skip, lid | 163 | Sponge swab | 2,613,760 |
| 35 | Pantry (staff & patient) | Drinks Cooler, pre-Flush | 53 | Wastewater | 1,032,506 |
| 41 | Drug Prep Room | Sink drain, U bend exterior | 44 | Sponge swab | 790,760 |
| 60 | Workstation on wheels | Keyboard and Mouse | 34 | Sponge swab | 864,837 |
| 71 | Bay (Beds 1-4) | Bathroom HWB Drain | 74 | Stick swab | 1,388,557 |
| 74 | Bay (Beds 1-4) | Bathroom toilet bowl | 48 | Stick swab | 1,037,570 |
| 75 | Bay (Beds 1-4) | Bathroom toilet floor | 22 | Sponge swab | 581,158 |
| 84 | Bay (Beds 5-8) | Bay Medication Cupboard | 98 | Sponge swab | 1,597,559 |
| 97 | Bed 9 | Bathroom HWB Drain | 74 | Stick swab | 1,201,404 |
| 102 | Bed 9 | Bathroom toilet bowl | 118 | Stick swab | 2,036,935 |
| 142 | Bay (Beds 15-18) | Bathroom toilet bowl | 70 | Stick swab | 1,315,258 |
| 158 | Bay (Beds 21-24) | Bathroom toilet bowl | 145 | Stick swab | 2,429,559 |
| 176 | Staff toilet | Staff toilet HWB DWT | 290 | Wastewater | 4,156,799 |
| 177 | Staff toilet | Staff toilet HWB DWT | 75 | Sponge swab | 1,279,241 |
| 178 | Pantry (Staff & patient) | Sink DWT | 93 | Wastewater | 1,818,705 |
| 179 | Pantry (Staff & patient) | Sink DWT | 56 | Sponge swab | 1,284,616 |
| 182 | Room 33 | Office HWB DWT | 328 | Wastewater | 4,703,504 |
| 183 | Room 33 | Office HWB DWT | 177 | Sponge swab | 2,537,593 |
| 184 | Bay (Beds 1-4) | HWB DWT | 63 | Wastewater | 1,083,094 |
| 185 | Bay (Beds 1-4) | HWB DWT | 39 | Sponge swab | 654,450 |
| 186 | Bay (Beds 5-8) | HWB DWT | 440 | Wastewater | 6,622,675 |
| 187 | Bay (Beds 5-8) | HWB DWT | 126 | Sponge swab | 2,021,118 |
| 190 | Bed 9 | HWB DWT | 263 | Wastewater | 6,456,693 |
| 191 | Bed 9 | HWB DWT | 111 | Sponge swab | 2,078,635 |
| 194 | Bed 10 | HWB DWT | 51 | Wastewater | 795,058 |
| 195 | Bed 10 | HWB DWT | 88 | Sponge swab | 1,573,130 |
| 198 | Bay (Beds 15-18) | HWB DWT | 171 | Wastewater | 11,334,805 |
| 199 | Bay (Beds 15-18) | HWB DWT | 53 | Sponge swab | 9,700,533 |
| 202 | Bed 19 | HWB DWT | 160 | Wastewater | 2,521,155 |
| 203 | Bed 19 | HWB DWT | 41 | Sponge swab | 13,997,999 |
| 206 | Bay (Beds 29-32) | HWB DWT | 255 | Wastewater | 4,058,517 |
| 207 | Bay (Beds 29-32) | HWB DWT | 37 | Sponge swab | 789,087 |
| 208 | Bay (Beds 29-32) | Bathroom HWB DWT | 448 | Wastewater | 12,313,813 |
| 209 | Bay (Beds 29-32) | Bathroom HWB DWT | 14 | Sponge swab | 210,049 |
| 212 | Bed 20 | HWB DWT | 409 | Wastewater | 6,070,475 |
| 213 | Bed 20 | HWB DWT | 43 | Sponge swab | 30,494,541 |

### 3.3 AMR genes

#### 3.3.1 Clinical isolates

Of the clinical isolates, *bla_OXA-48_* was identified in 2 of the total 28 (7.1%) isolates (one *C. portucalensis* and one *K. pneumoniae*). *bla_NDM-1_* was identified in nine (32.1%) clinical isolates (one *C. portucalensis*, five *E. hormaechei*, one *E. coli* and two *K. pneumoniae*). *bla*_OXA_ genes on the CARBA-5 panel were found in 84% of clinical isolates and *bla*_NDM_ genes on the CARBA-5 panel were found in 76% of clinical isolates, see Table 7. See supplementary materials S4 for full list of genes found.

**Table 7.** Clinical isolates with *bla_OXA-48_*, at least one *bla_OXA_* (on the CARBA5 panel), total number of isolates with any *bla_OXA_* gene, *bla_NDM-1_*, at least one *bla_NDM_* (on the CARBA5 panel) and total number of isolates with any *bla_NDM_* gene.

| Species | Total isolates | *bla*<sub>OXA-48</sub> | *bla*<sub>OXA</sub> | Any *bla*<sub>OXA</sub> | *bla*<sub>NDM-1</sub> | *bla*<sub>NDM</sub> | Any *bla*<sub>NDM</sub> |
| --- | --- | --- | --- | --- | --- | --- | --- |
| *C. portucalensis* | 1 | 1 (100%) | 1 (100%) | 1 (100%) | 1 (100%) | 1 (100%) | 1 (100%) |
| *E. hormaechei* | 5 | 0 (0.0%) | 0 (0.0%) | 0 (0.0%) | 5 (100%) | 5 (100%) | 5 (100%) |
| *E. coli* | 9 | 0 (0.0%) | 8 (89%) | 8 (89%) | 1 (11%) | 1 (11%) | 1 (11%) |
| *K. pneumoniae* | 10 | 1 (10%) | 9 (90%) | 9 (90%) | 2 (20%) | 10 (100%) | 10 (100%) |
| Total | 25 | 2 (8.0%) | 21 (84%) | 21 (84%) | 9 (32%) | 19 (76%) | 19 (76%) |

**Table 8.**
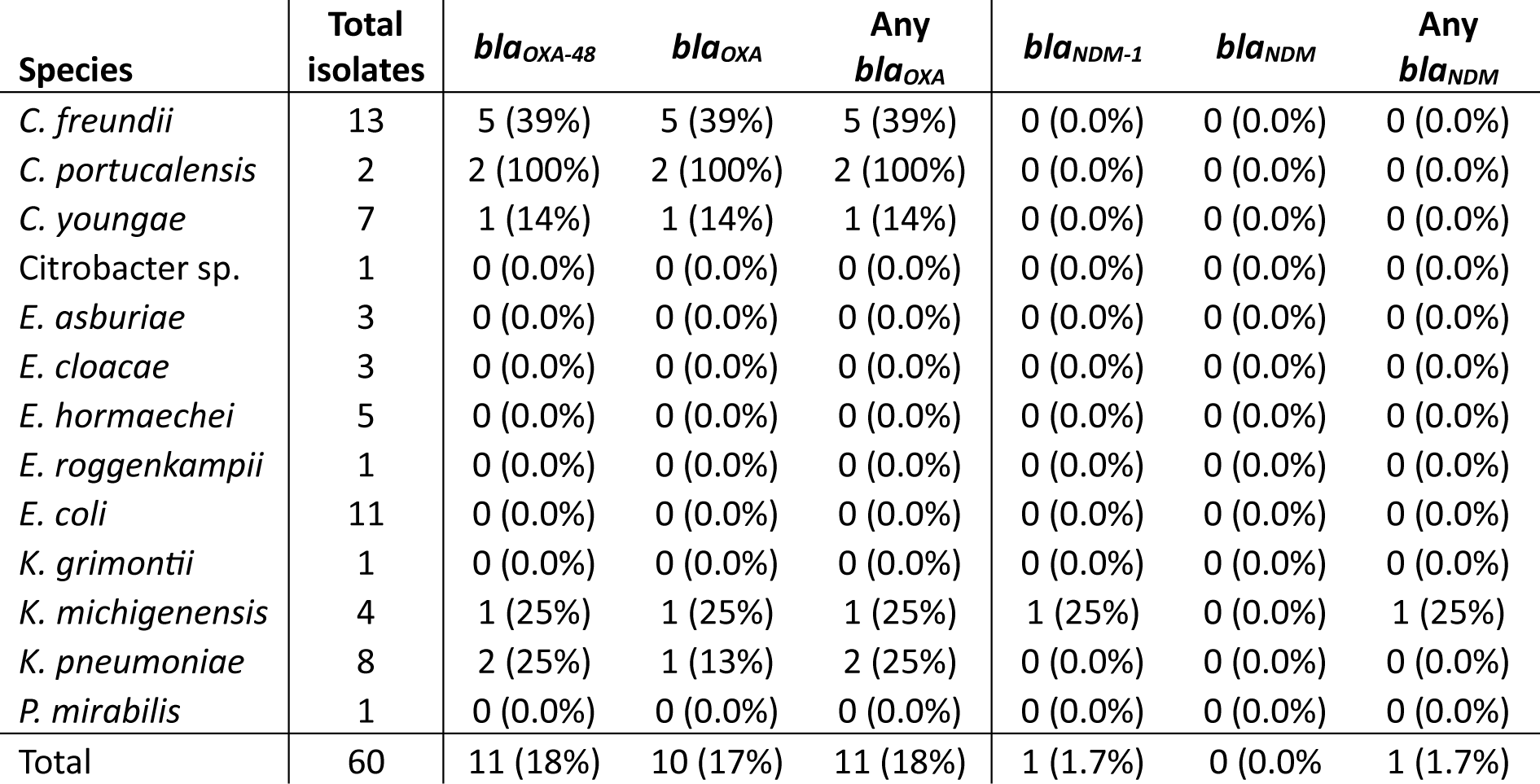
Environmental isolates with *bla_OXA-48_*, at least one *bla_OXA_* (on the CARBA5 panel), total number of isolates with any *bla_OXA_* gene, *bla_NDM-1_*, at least one *bla_NDM_* (on the CARBA5 panel) and total number of isolates with any *bla_NDM_* gene.

| Species | Total isolates | *bla*<sub>OXA-48</sub> | *bla*<sub>OXA</sub> | Any *bla*<sub>OXA</sub> | *bla*<sub>NDM-1</sub> | *bla*<sub>NDM</sub> | Any *bla*<sub>NDM</sub> |
| --- | --- | --- | --- | --- | --- | --- | --- |
| *C. freundii* | 13 | 5 (39%) | 5 (39%) | 5 (39%) | 0 (0.0%) | 0 (0.0%) | 0 (0.0%) |
| *C. portucalensis* | 2 | 2 (100%) | 2 (100%) | 2 (100%) | 0 (0.0%) | 0 (0.0%) | 0 (0.0%) |
| *C. youngae* | 7 | 1 (14%) | 1 (14%) | 1 (14%) | 0 (0.0%) | 0 (0.0%) | 0 (0.0%) |
| Citrobacter sp. | 1 | 0 (0.0%) | 0 (0.0%) | 0 (0.0%) | 0 (0.0%) | 0 (0.0%) | 0 (0.0%) |
| *E. asburiae* | 3 | 0 (0.0%) | 0 (0.0%) | 0 (0.0%) | 0 (0.0%) | 0 (0.0%) | 0 (0.0%) |
| *E. cloacae* | 3 | 0 (0.0%) | 0 (0.0%) | 0 (0.0%) | 0 (0.0%) | 0 (0.0%) | 0 (0.0%) |
| *E. hormaechei* | 5 | 0 (0.0%) | 0 (0.0%) | 0 (0.0%) | 0 (0.0%) | 0 (0.0%) | 0 (0.0%) |
| *E. roggkampii* | 1 | 0 (0.0%) | 0 (0.0%) | 0 (0.0%) | 0 (0.0%) | 0 (0.0%) | 0 (0.0%) |
| *E. coli* | 11 | 0 (0.0%) | 0 (0.0%) | 0 (0.0%) | 0 (0.0%) | 0 (0.0%) | 0 (0.0%) |
| *K. grimontii* | 1 | 0 (0.0%) | 0 (0.0%) | 0 (0.0%) | 0 (0.0%) | 0 (0.0%) | 0 (0.0%) |
| *K. michigenensis* | 4 | 1 (25%) | 1 (25%) | 1 (25%) | 1 (25%) | 0 (0.0%) | 1 (25%) |
| *K. pneumoniae* | 8 | 2 (25%) | 1 (13%) | 2 (25%) | 0 (0.0%) | 0 (0.0%) | 0 (0.0%) |
| *P. mirabilis* | 1 | 0 (0.0%) | 0 (0.0%) | 0 (0.0%) | 0 (0.0%) | 0 (0.0%) | 0 (0.0%) |
| Total | 60 | 11 (18%) | 10 (17%) | 11 (18%) | 1 (1.7%) | 0 (0.0%) | 1 (1.7%) |

#### 3.3.2 Environmental isolates

Of the environmental isolates, *bla_OXA-48_* was identified in 11 of the total 60 (18%) isolates (See Table 88). *bla_NDM-1_* was identified in one (1.7%) environmental isolate, a *K. michigenensis*. See supplementary materials S5 for full list of genes found.

#### 3.3.3 Metagenomic environmental samples

*bla_OXA-48_* was identified in 1/36 (2.8%) metagenomic environmental sample, and three other *bla_OXA_* genes on the CARBA5 panel (*bla_OXA-204_*, *bla_OXA-370_* and *bla_OXA-515_*) were found together in 1/36 (2.8%) other metagenomic environmental sample. No *bla_NDM_* genes were identified in any of the 36 metagenomic environmental samples.

### 3.4 Plasmids

#### 3.4.1 Clinical isolates

The clinical isolates had a mean number of 5.6 plasmids (SD = 2.3) and a total of 20 different plasmids were identified. Table 9 describes the plasmid types and species they were found in. Col440II was the only plasmid found across all clinical isolate species. IncFIB(AP001918) was the most commonly identified plasmid.

**Table 9.** Plasmid types found in percentage of clinical isolates. Col440II was the only plasmid found across all clinical isolate species. IncFIB(AP001918) was the most commonly identified plasmid.

| Target plasmid | *C. portucalensis* | *E. hormaechei* | *E. coli* | *K. pneumoniae* | Total no. isolates found in |
| --- | --- | --- | --- | --- | --- |
| Col(pHAD28) | 0 (0.0%) | 5 (100%) | 0 (0.0%) | 0 (0.0%) | 5 (17.9%) |
| Col156 | 0 (0.0%) | 0 (0.0%) | 9 (90.0%) | 0 (0.0%) | 9 (32.1%) |
| Col440I | 1 (100%) | 0 (0.0%) | 2 (20.0%) | 12 (100%) | 15 (53.6%) |
| Col440II | 1 (100%) | 1 (20.0%) | 2 (20.0%) | 12 (100%) | 16 (57.1%) |
| ColKP3 | 1 (100%) | 0 (0.0%) | 1 (10.0%) | 10 (83.3%) | 12 (42.9%) |
| IncC | 1 (100%) | 0 (0.0%) | 0 (0.0%) | 0 (0.0%) | 1 (3.6%) |
| IncFIA | 0 (0.0%) | 0 (0.0%) | 9 (90.0%) | 11 (91.7%) | 20 (71.4%) |
| IncFIB(AP001918) | 0 (0.0%) | 0 (0.0%) | 9 (90.0%) | 9 (75.0%) | 18 (64.3%) |
| IncFIB(K) | 0 (0.0%) | 0 (0.0%) | 0 (0.0%) | 1 (8.3%) | 1 (3.6%) |
| IncFIB(pQil) | 0 (0.0%) | 0 (0.0%) | 0 (0.0%) | 11 (91.7%) | 11 (39.3%) |
| IncFII | 0 (0.0%) | 0 (0.0%) | 9 (90.0%) | 4 (33.3%) | 13 (46.4%) |
| IncFII(K) | 0 (0.0%) | 0 (0.0%) | 0 (0.0%) | 11 (91.7%) | 11 (39.3%) |
| IncHI1A(NDM-CIT) | 1 (100%) | 0 (0.0%) | 0 (0.0%) | 0 (0.0%) | 1 (3.6%) |
| IncHI1B(pNDM-CIT) | 1 (100%) | 0 (0.0%) | 0 (0.0%) | 0 (0.0%) | 1 (3.6%) |
| IncHI2 | 0 (0.0%) | 0 (0.0%) | 1 (10.0%) | 1 (8.3%) | 2 (7.1%) |
| IncHI2A | 0 (0.0%) | 0 (0.0%) | 1 (10.0%) | 1 (8.3%) | 2 (7.2%) |
| IncL | 1 (100%) | 0 (0.0%) | 0 (0.0%) | 0 (0.0%) | 1 (3.6%) |
| IncR | 0 (0.0%) | 0 (0.0%) | 0 (0.0%) | 1 (8.3%) | 1 (3.6%) |
| IncX3 | 0 (0.0%) | 5 (100%) | 0 (0.0%) | 11 (91.7%) | 16 (57.1%) |
| p0111 | 0 (0.0%) | 0 (0.0%) | 1 (10.0%) | 0 (0.0%) | 1 (3.6%) |

#### 3.4.2 Environmental isolates

The environmental isolates had a mean number of 4.3 plasmids (SD = 2.3) and a total of 42 different plasmids were identified. Supplementary materials S6 describes the plasmids and species they were found in. Col(IRGK) was the most commonly identified plasmid in 35/60 (59.3%) isolates, no plasmid was ubiquitous across all environmental isolate species.

#### 3.4.3 Metagenomic environmental samples

Plasmids for Enterobacterales were found in 24/36 (66.6%) environmental metagenomic samples (see Supplementary materials S7). The mean number of plasmids was 3.1 (SD = 3.6).

### 3.5 Metagenomic species

When species were analysed in the metagenomic samples, reads mapping to the Enterobacterales order were found in 64%-75% of sites (see Table 10). When analysed, there was no significant difference in the number of reads that mapped to *Citrobacter*, *Enterobacter*, *Escherichia* or *Klebsiella* genera, using One Way ANOVA and Tukey’s multiple comparison test.

**Table 10.** Number of metagenomic samples sites for which reads mapped to Enterobacterales genus sequences.

| Enterobacterales genera | Number of swab locations found in | Mean number of reads (SD) |
| --- | --- | --- |
| *Citrobacter* | 26 (72%) | 1,229,432 (4,492,888) |
| *Enterobacter* | 23 (64%) | 122,971 (316,369) |
| *Escherichia* | 27 (75%) | 171,588 (749,207) |
| *Klebsiella* | 26 (72%) | 47,032 (71,755) |

### 3.6 Transmission and patient metadata

One large cluster of *C. freundii* isolates that mapped to strain N16-03880 were all from the same room (beds 29-32) but came from both the bay HWB DWT as well as the bathroom HWB DWT samples, suggesting cross-contamination of the two sinks (see Figure *1A*, *C. freundii* cluster #1). These, along with a toilet bowl swab from beds 15-18 were the only *C. freundii* found to harbour *bla*_OXA-48_ genes (see Figure 1A). All of the *C. youngae* environmental isolates mapped to strain CF10 (see Figure 1B, *C. youngae* cluster #1). The *C. portucalensis* isolates all mapped to different strains. The clinical isolate (patient Z1) mapped mainly (85% query coverage) to strain PNUCL1, and the environmental isolates mapped to FDAARGOS_617 and SWHIN_111. All harboured *bla*_OXA_ genes, and the patient isolate additionally harboured *bla_NDM_* genes (see Figure 1C).

**Figure 1.**
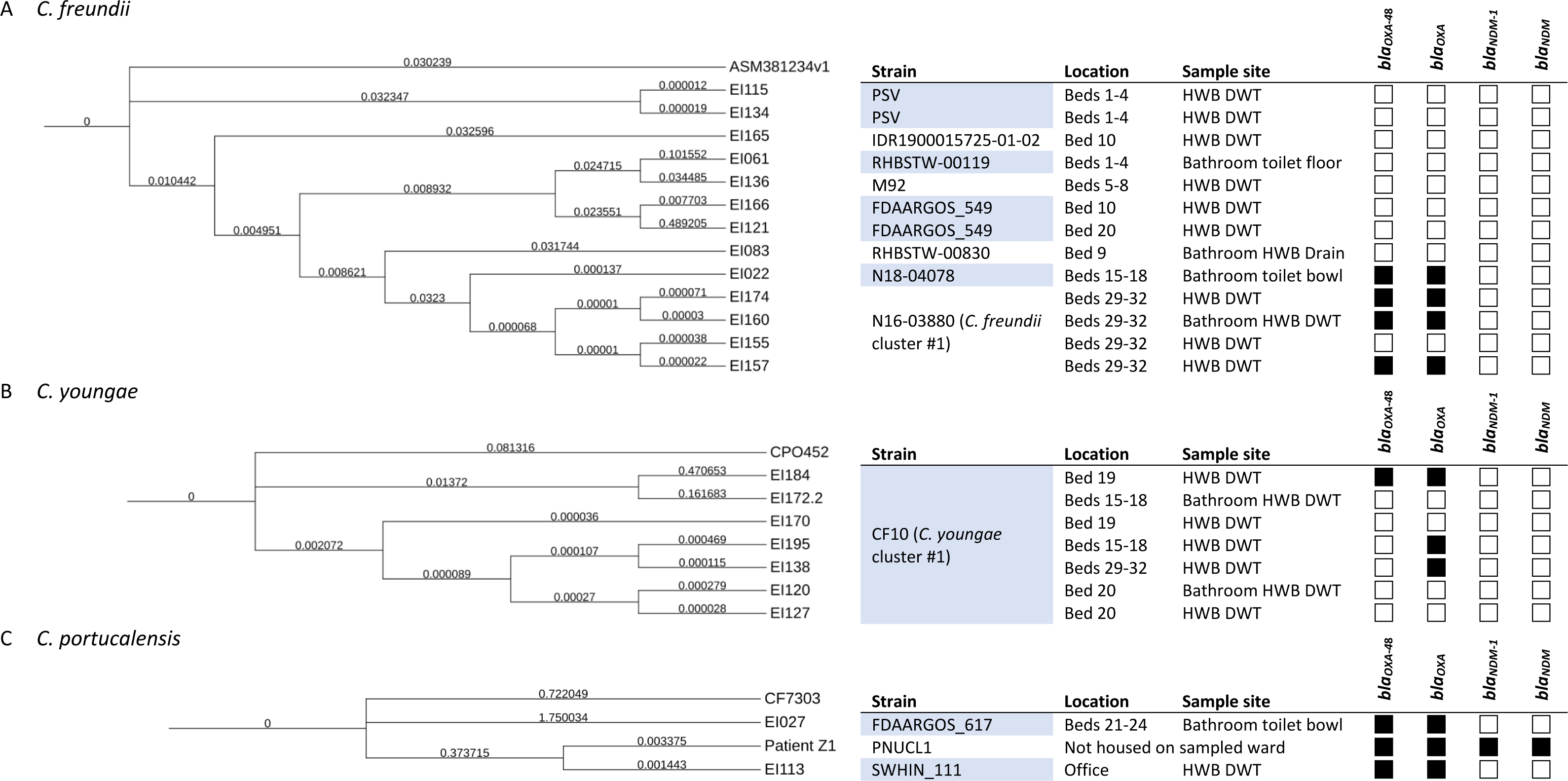
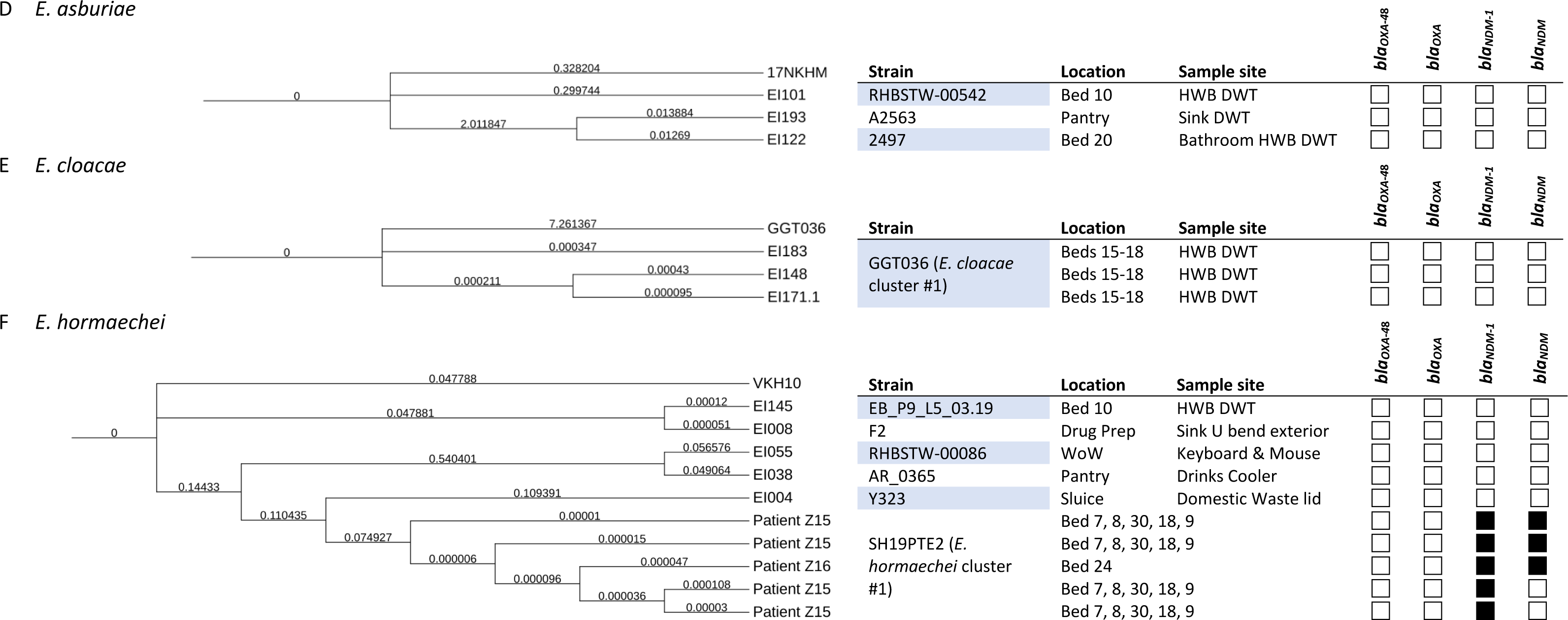
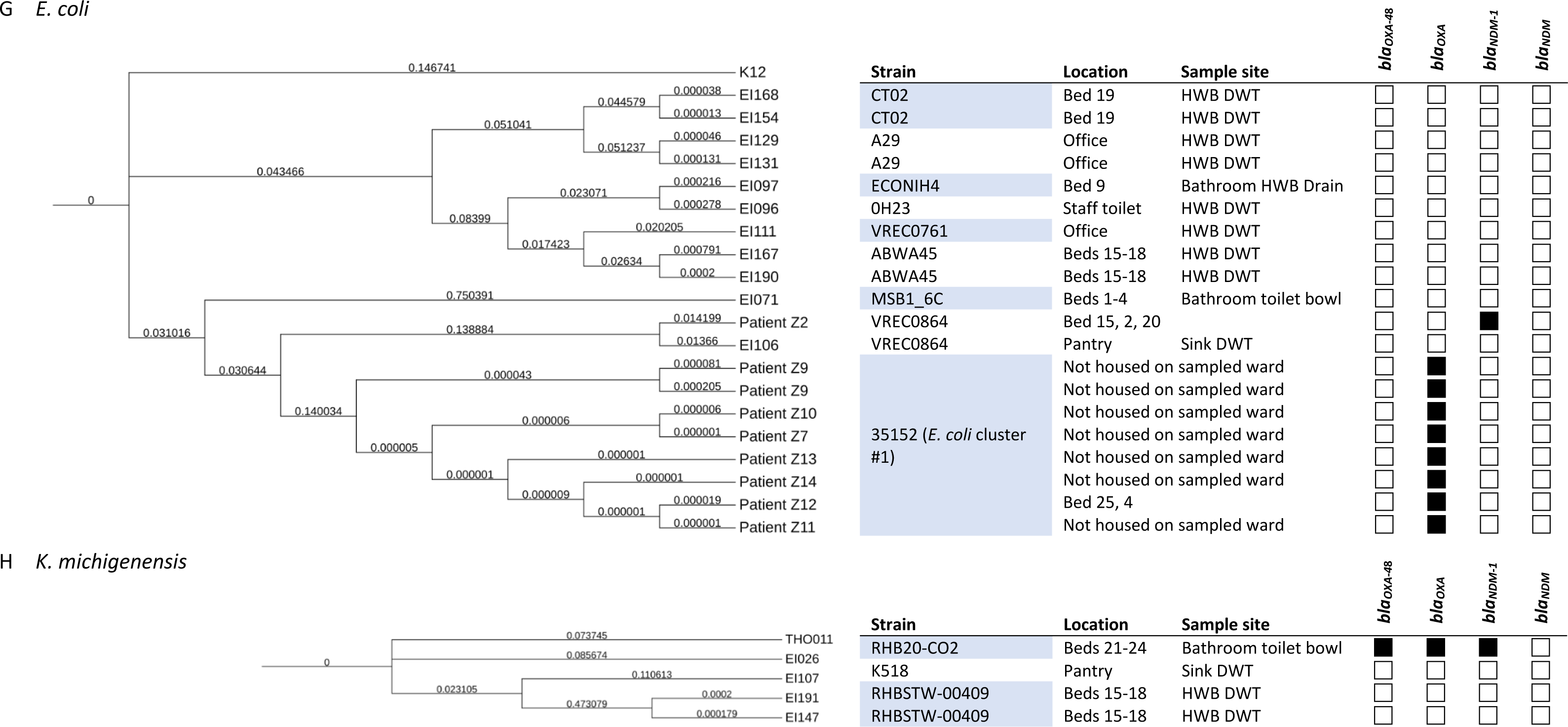
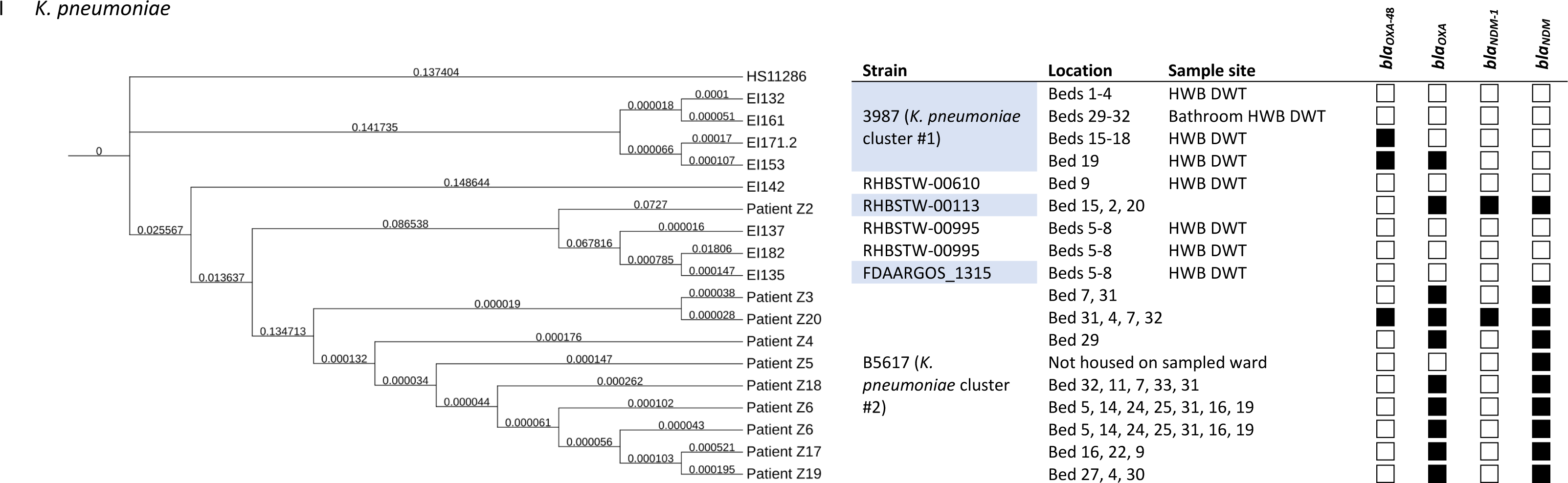
Phylogram of CRE species isolated in this study and metadata including location (of either sample collection for environmental isolates, or beds inhabited for clinical isolates), strain and presence or absence of *bla*_OXA_ and *bla*_NDM_ genes. A) C*. freundii*, B) *C. portucalensis*, C) C*. youngae*, D) *E. asburiae*, E) *E. cloacae*, F) *E. hormaechei*, G) *E. coli*, H) *K. michigenensis* and I) *K. pneumoniae*. Branch lengths indicate number of substitutions divided by the length of the genome sequence. HWB = hand wash basin, DWT = drain waste trap, WoW = workstation on wheels. *bla_OXA_* = *bla*_OXA_ gene on CARBA-5 panel, *bla_NDM_* = *bla*_NDM_ gene on CARBA5 panel, EI = Environmental isolate. The top branch of each phylogram is the reference genome (e.g. HS11286 for *K. pneumoniae*), against which the clinical and environmental isolates were mapped.

The *E. asburiae* environmental isolates all mapped to different strains: A2563, 2497 and RHBSTW-00542 (see Figure 1D). The *E. cloacae* environmental isolates all mapped to strain GGT036. Neither species were found to harbour any *bla*_OXA_ or *bla_NDM_* genes (see Figure 1E). All of the *E. hormaechei* clinical isolates mapped to strain SH19PTE2 and were found to harbour *bla_NDM_* genes, whereas the environmental isolates each mapped to a different strain: Y323, F2, AR_0365 and RHBSTW-00086, none of which harboured either *bla*_OXA_ or *bla_NDM_* genes (see Figure 1F).

Except patient Z2, all patient *E. coli* isolates mapped most closely to strain 035152. The 035152 isolates had a similar AMR profile, with all (9/9) harbouring *bla_OXA-181_* and 7/9 harbouring *bla_OXA-484_*. Patient Z2, which mapped most closely to *E. coli* VREC0864, clustered with EI106, an environmental isolate from a water sample from the pantry sink’s drain waste trap, which also mapped most closely to VREC0864 (see Figure 1G, *E. coli* cluster #1).

The environmental isolates that mapped to *K. michigenensis* mapped most closely to strains RHB20-CO2, K518 and RHBSTW-00409 (two isolates from the same HWB DWT in beds 15-18). Only one was found to harbour *bla*_OXA_ and *bla_NDM_* genes (see Figure *1*H). Except Z5 (strain RHBSTW-00113), all of the *K. pneumoniae* clinical isolates mapped most closely to strain B5617 (see Figure 1I, *K. pneumoniae* cluster #2). Patient isolates Z3, Z4, Z5, Z6, Z17, Z18 and Z19 (all strain B5617) showed a similar AMR profile, all harbouring *bla_OXA-181_, bla_OXA-232_* and *bla_OXA-484_*, and *bla_NDM-5_*. There was a cluster of environmental isolates that mapped most closely to *K. pneumoniae* strain 3987, all isolated from hand wash basin drain waste traps, but in differing patient bed areas around the ward (see Figure 1I, *K. pneumoniae* cluster #1).

9/20 (45%) patients were housed on ward 7D during their time as an inpatient and 9/9 (100%) patients had stayed in bay (beds 15-18), bay (beds 29-32) or in one of the single-occupancy bed rooms, each of which were noted to have had higher numbers of Enterobacterales isolated from the environmental samples. 4/9 (44%) of these patients had been housed in more than one of the bays, one patient had been on both bay (beds 15-18), bay (beds 29-32) and bay 19.

## 4 Discussion

There was evidence for the potential for patient-patient transmission for *E. hormaechei*, *E. coli* and *K. pneumoniae*. This conclusion is supported by the patient spatio-temporal data collected. This study also identified environment-patient transmission. The *E. coli* isolate from patient Z2 clustered with EI106, an environmental isolate from a wastewater sample taken from the communal pantry sink’s DWT, although they did not share the same plasmids. Whilst the clinical isolate was found to harbour the *bla*_NDM-1_ gene, no *bla*_OXA_ or *bla*_NDM_ genes were found in the environmental isolate. This may have been a loss or gain of function; bacteria found within the environment are less likely to encounter antibiotics or their residues compared with those in a hospitalised patient, that isolate may have lost the plasmid containing these resistance genes, conversely, they may gain plasmids from commensal organisms within the host [43].

It was noted that Enterobacterales were commonly isolated from environmental samples in bay (beds 15-18), bay (beds 29-32) and the single-occupancy bays. All the patients in this study located on ward 7D had been housed in at least one of these bays and 44% had been housed in more than one of these bays, suggesting potential host-related reservoirs. One report suggests that patients are on average 73% more likely to acquire a HAI if the patient previously occupying their room was colonised or infected, [44]. This suggests enhanced location-specific IPC would be beneficial when colonised or infected patients have been identified, and that isolation of these patients may not be enough. Enterobacterales were also isolated from environmental samples in non-clinical areas such as the shared-use pantry and the staff office. Studies have shown that HCWs can be colonised when handling patients and infected materials, it’s also possible that colonised patients or HCWs may have caused reservoirs in the pantry due to transmission of organisms via the faecal-oral route [45].

Even with comprehensive patient metadata, such as bed movements, it can be difficult to confidently infer transmission of a clonal isolate. Environmental isolates are especially complex, as the bacteria may have been present for long periods, for example by forming hard to remove biofilms in U-bends, which is in itself difficult to monitor and identify provenance [46]. For human pathogens, the ward environment may not be optimal for growth, so their doubling time may be slower. Genetic-relatedness cutoffs to determine phylogeny tend to be calculated depending on the sample number, type and environments, as well as the species, as some have faster molecular clocks than others. These data are often missing for isolates extracted from the environment. [47].

Whilst all clinical isolates tested positive for OXA-48-like variants using the CARBA-5 test, only 84% of the clinical isolates were found to harbour *bla*_OXA_ genes that the CARBA-5 panel tested for. Studies have shown the specificity of the CARBA-5 test to vary from 96% (from blood cultures) to 100% from isolates and rectal swabs [48], [49], [50], [51]. The clinical sites that the swabs were taken from in this study varied and the majority came from CRE screens, usually rectal, rather than sites of infection. It is possible that the CARBA-5 panel picked up *bla*_OXA_ genes from other colonising species present in CRE screening samples, as these were not sterile sites. Most of the clinical and environmental isolates found to harbour the *bla*_OXA-48_ gene also had other *bla*_OXA_ genes present, as well as other resistance genes, such as *bla*_CTX_ and *bla*_SHV_. These are often found together on the same plasmids and are likely to be transferred between bacteria collectively [4]. Indeed, all of the *C. portucalensis* isolates identified in this study were found to harbour *bla*_OXA_ genes, with the clinical isolate also containing *bla*_NDM-1_.

Whilst Col440II, which does not carry resistance or virulence genes, was the only plasmid found across all clinical isolate species. IncFIB(AP001918), the most commonly identified plasmid, is linked with resistance to several antimicrobial classes, including β-lactams, aminoglycosides, sulfonamides and tetracyclines, but was less commonly identified in environmental isolates [52], [53]. The prevalence of IncFIB(AP001918) in clinical samples suggests that genomic analysis of plasmids as well as isolates is important for enhanced IPC surveillance.

In this study, the initial MALDI-TOF speciation for *C. freundii*, *E. cloacae* and *K. oxytoca* isolates did not all match with the WGS speciation. Both methods were more consistent when comparing environmental rather than clinical isolates. All three species complexes contain multiple, closely related species, which can make it difficult to fully resolve using conventional methods*. C. portucalensis* is a relatively newly described clinical pathogen but has the capacity to harbour and transmit AMR genes, thus identifying it to species level may be important in the future [54].

The use of genomic analysis in enhanced outbreak surveillance technologies provides greater detail on the potential transmission of MDROs and their associations with patients, HCWs and the ward Environment. Platforms such as ONT shows promise, especially for use in low- and middle-income countries, as long read sequencing enables read lengths of thousands, rather than hundreds, of base pairs which is especially useful when resolving speciation in metagenomic samples.

## 5 Conclusions

Understanding the resistance genes, plasmids and sequencing types present in an environment can provide greater resolution than phenotypic and other molecular methods, helping to identify targeted IPC interventions in outbreak situations. As a result of the evidence from this study highlighting the presence of CREs in wastewater, the hospital estates team has since replaced all of the sink U-bends on the sampled ward, as well as reviewed and revised ward IPC practices to reduce potential transmission risks. Due to the number of colonised patients found, this study also recommends the use of more widespread CRE screening for hospitalised patients, to enable interventions to reduce the risks from human and environmental reservoirs and therefore reduce risks to vulnerable patients.

Overview:

- Putative patient-patient and patient-ward transmission was identified, utilising WGS and patient metadata
- CREs were more commonly isolated from wastewater samples than either stick swabs or sponge swabs
- All patients in this study tested positive for OXA-48-like carbapenemase variants using the NG-Test® CARBA-5 rapid diagnostic test, 84% of patient isolates harboured a *bla_OXA_* gene present on the CARBA-5 panel
- *bla*_OXA_ and *bla_NDM_* genes were identified in fewer environmental CRE isolates, compared with clinical isolates, suggesting either different populations, or a loss/gain of plasmids or genes

## Ethical approval

This diagnostic service evaluation was undertaken at the request of the Division of Nursing in support of the Infection Prevention and Control team as part of enhanced standard of care practices for IPC outbreak investigation. This project is under HRA approval for ELCID and registered with Health Services London. IRAS project ID: 283831, National HRA REC reference: 20/HRA/4928, Local REC: RFL R&D ref: 134895, HSL Project number: 1529

## Data sharing

All genomic data produced in the present work are contained in the manuscript or available online under ENA bioproject number PRJEB76684. For patient and ward metadata, please contact the authors.

## Data Availability

All data produced in the present work are contained in the manuscript or available online under ENA bioproject number PRJEB76684. For patient and ward metadata, please contact the authors.

## Abbreviations

AMR: Antimicrobial resistance
AST: Antibiotic sensitivity test
CRE: Carbapenem-resistant Enterobacterales
DWT: Drain waste trap
HCAI: Healthcare associated infection
HWB: Hand wash basin
IPC: Infection prevention and control
MDRO: Multi-drug resistant organism
NDM: New Delhi metallo-beta lactamase
ONT: Oxford Nanopore Technologies
RDT: Rapid diagnostic test
SD: Standard deviation
WoW: Workstation on wheels

## References

[1] World Health Organization, “Guidelines for the prevention and control of carbapenem-resistant Enterobacteriaceae, Acinetobacter baumannii and Pseudomonas aeruginosa in health care facilities,” 2017. [Online]. Available: http://apps.who.int/bookorders.

[2] M. C. Mills and J. Lee, “The threat of carbapenem-resistant bacteria in the environment: Evidence of widespread contamination of reservoirs at a global scale,” Environmental Pollution, vol. 255. Elsevier Ltd, Dec. 01, 2019. doi: 10.1016/j.envpol.2019.113143.

[3] T. Tängdén and C. G. Giske, “Global dissemination of extensively drug-resistant carbapenemase-producing Enterobacteriaceae: Clinical perspectives on detection, treatment and infection control,” Journal of Internal Medicine, vol. 277, no. 5. Blackwell Publishing Ltd, pp. 501–512, May 01, 2015. doi: 10.1111/joim.12342.

[4] K. Kopotsa, J. Osei Sekyere, and N. M. Mbelle, “Plasmid evolution in carbapenemase-producing Enterobacteriaceae: a review,” Annals of the New York Academy of Sciences, vol. 1457, no. 1. Blackwell Publishing Inc., pp. 61–91, Dec. 01, 2019. doi: 10.1111/nyas.14223.

[5] P. Nordmann et al., “Identification and screening of carbapenemase-producing Enterobacteriaceae,” Clinical Microbiology and Infection, vol. 18, no. 5. Blackwell Publishing Ltd, pp. 432–438, May 01, 2012. doi: 10.1111/j.1469-0691.2012.03815.x.

[6] N. Gupta, B. M. Limbago, J. B. Patel, and A. J. Kallen, “Carbapenem-resistant enterobacteriaceae: Epidemiology and prevention,” Clinical Infectious Diseases, vol. 53, no. 1. pp. 60–67, Jul. 01, 2011. doi: 10.1093/cid/cir202.

[7] P. A. Apanga et al., “Carbapenem-resistant Enterobacteriaceae in sink drains of 40 healthcare facilities in Sindh, Pakistan: A cross-sectional study,” PLoS One, vol. 17, no. 2 February, Feb. 2022, doi: 10.1371/journal.pone.0263297.

[8] C. A. Islam et al., “Environmental Spread of New Delhi Metallo-Lactamase-1-Producing Multidrug-Resistant Bacteria in Dhaka, Bangladesh Environmental spread of New Delhi metallo-β-lactamase-1-producing multidrug-resistant bacteria in Dhaka,” 2017. [Online]. Available: 10

[9] K. Zurfluh et al., “Wastewater is a reservoir for clinically relevant carbapenemase-and 16s rRNA methylase-producing Enterobacteriaceae,” Int J Antimicrob Agents, vol. 50, no. 3, pp. 436–440, Sep. 2017, doi: 10.1016/j.ijantimicag.2017.04.017.

[10] K. Zenati, A. Touati, S. Bakour, F. Sahli, and J. M. Rolain, “Characterization of NDM-1-and OXA-23-producing Acinetobacter baumannii isolates from inanimate surfaces in a hospital environment in Algeria,” Journal of Hospital Infection, vol. 92, no. 1, pp. 19–26, Jan. 2016, doi: 10.1016/j.jhin.2015.09.020.

[11] M. Perry et al., “Antimicrobial resistance in hospital wastewater in Scotland: a cross-sectional metagenomics study,” The Lancet, vol. 394, p. S1, Nov. 2019, doi: 10.1016/s0140-6736(19)32798-9.

[12] Department of Health and Social Care, “Sewage in water: a growing public health problem.” Accessed: Jun. 23, 2024. [Online]. Available: https://www.gov.uk/government/news/sewage-in-water-a-growing-public-health-problem

[13] U. Klümper et al., “Environmental microbiome diversity and stability is a barrier to antimicrobial resistance gene accumulation,” Commun Biol, vol. 7, no. 1, p. 706, Jun. 2024, doi: 10.1038/s42003-024-06338-8.

[14] M. J. Struelens, C. Ludden, G. Werner, V. Sintchenko, P. Jokelainen, and M. Ip, “Real-time genomic surveillance for enhanced control of infectious diseases and antimicrobial resistance,” Frontiers in Science, vol. 2, Apr. 2024, doi: 10.3389/fsci.2024.1298248.

[15] Clinical and Laboratory Standards Institute, “Performance Standards for Antimicrobial Susceptibility Testing,” in CLSI supplement M100, 31st ed., 2021.

[16] Z. Chen et al., “Global landscape of SARS-CoV-2 genomic surveillance and data sharing,” Nat Genet, vol. 54, no. 4, pp. 499–507, Apr. 2022, doi: 10.1038/s41588-022-01033-y.

[17] M. Boolchandani, A. W. D’Souza, and G. Dantas, “Sequencing-based methods and resources to study antimicrobial resistance,” Nature Reviews Genetics, vol. 20, no. 6. Nature Publishing Group, pp. 356–370, Jun. 01, 2019. doi: 10.1038/s41576-019-0108-4.

[18] European Committee on Antimicrobial Susceptibility Testing, “Breakpoint tables for interpretation of MICs and zone diameters (v12),” 2022.

[19] Hardy Diagnostics, “NG-Test CARBA 5,” 2021.

[20] S. Ali and A. P. R. Wilson, “Effect of poly-hexamethylene biguanide hydrochloride (PHMB) treated non-sterile medical gloves upon the transmission of Streptococcus pyogenes, carbapenem-resistant E. coli, MRSA and Klebsiella pneumoniae from contact surfaces,” BMC Infect Dis, vol. 17, no. 1, Aug. 2017, doi: 10.1186/s12879-017-2661-9.

[21] Ö. Yetiş, S. Ali, K. Karia, P. Bassett, and P. Wilson, “Enhanced monitoring of healthcare shower water in augmented and non-augmented care wards showing persistence of Pseudomonas aeruginosa despite remediation work,” J Med Microbiol, vol. 72, no. 5, 2023, doi: 10.1099/jmm.0.001698.

[22] Qiagen, “Sample to Insight DNeasy ® Blood & Tissue Handbook,” 2023.

[23] Zymo Research, “ZymoBIOMICS^TM^ DNA Miniprep Kit.” Accessed: Aug. 22, 2023. [Online]. Available: https://files.zymoresearch.com/protocols/_d4300t_d4300_d4304_zymobiomics_dna_miniprep_kit.pdf

[24] Zymo Research, “ZymoBIOMICS® Microbial Community Standard.” Accessed: Aug. 22, 2023. [Online]. Available: https://files.zymoresearch.com/protocols/_d6300_zymobiomics_microbial_community_standard.pdf

[25] Oxford Nanopore Technologies, “Rapid sequencing gDNA - barcoding (SQK-RBK110.96).” Accessed: Dec. 12, 2022. [Online]. Available: https://community.nanoporetech.com/docs/prepare/library_prep_protocols/rapid-barcoding-kit-96-sqk-rbk110-96/v/rbk_9126_v110_revm_24mar2021

[26] Oxford Nanopore Technologies, “Rapid PCR Barcoding Kit (SQK-RPB004) protocol.” Accessed: Jun. 28, 2021. [Online]. Available: https://community.nanoporetech.com/protocols/rapid-pcr-barcoding/checklist_example.pdf

[27] Zymo Research, “ZymoBIOMICS^TM^ Microbial Community DNA Standard.” Accessed: Aug. 22, 2023. [Online]. Available: https://files.zymoresearch.com/protocols/_d6305_d6306_zymobiomics_microbial_community_dna_standard.pdf

[28] Babraham Bioinformatics, “FastQC.” Accessed: Jun. 28, 2021. [Online]. Available: https://www.bioinformatics.babraham.ac.uk/projects/fastqc/

[29] P. Ewels, M. Magnusson, S. Lundin, and M. Käller, “MultiQC: Summarize analysis results for multiple tools and samples in a single report,” Bioinformatics, vol. 32, no. 19, pp. 3047–3048, 2016, doi: 10.1093/bioinformatics/btw354.

[30] P. T. L. C. Clausen, F. M. Aarestrup, and O. Lund, “Rapid and precise alignment of raw reads against redundant databases with KMA,” BMC Bioinformatics, vol. 19, no. 1, pp. 1–8, 2018, doi: 10.1186/s12859-018-2336-6.

[31] P. T. L. C. Clausen, E. Zankari, F. M. Aarestrup, and O. Lund, “Benchmarking of methods for identification of antimicrobial resistance genes in bacterial whole genome data,” Journal of Antimicrobial Chemotherapy, vol. 71, no. 9, pp. 2484–2488, 2016, doi: 10.1093/jac/dkw184.

[32] H. Hasman et al., “Rapid whole-genome sequencing for detection and characterization of microorganisms directly from clinical samples,” J Clin Microbiol, vol. 52, no. 1, pp. 139–146, Jan. 2014, doi: 10.1128/JCM.02452-13.

[33] M. V. Larsen et al., “Benchmarking of methods for genomic taxonomy,” J Clin Microbiol, vol. 52, no. 5, pp. 1529–1539, 2014, doi: 10.1128/JCM.02981-13.

[34] A. Carattoli et al., “In Silico detection and typing of plasmids using plasmidfinder and plasmid multilocus sequence typing,” Antimicrob Agents Chemother, vol. 58, no. 7, pp. 3895–3903, 2014, doi: 10.1128/AAC.02412-14.

[35] H. Li, “Minimap2: Pairwise alignment for nucleotide sequences,” Bioinformatics, vol. 34, no. 18, pp. 3094–3100, 2018, doi: 10.1093/bioinformatics/bty191.

[36] H. Li et al., “The Sequence Alignment/Map format and SAMtools,” Bioinformatics, vol. 25, no. 16, pp. 2078–2079, 2009, doi: 10.1093/bioinformatics/btp352.

[37] T. Carver, S. R. Harris, M. Berriman, J. Parkhill, and J. A. McQuillan, “Artemis: An integrated platform for visualization and analysis of high-throughput sequence-based experimental data,” Bioinformatics, vol. 28, no. 4, pp. 464–469, 2012, doi: 10.1093/bioinformatics/btr703.

[38] T. J. Treangen, B. D. Ondov, S. Koren, and A. M. Phillippy, “The harvest suite for rapid core-genome alignment and visualization of thousands of intraspecific microbial genomes,” Genome Biol, vol. 15, no. 11, 2014, doi: 10.1186/s13059-014-0524-x.

[39] M. N. Price, P. S. Dehal, and A. P. Arkin, “FastTree 2 - Approximately maximum-likelihood trees for large alignments,” PLoS One, vol. 5, no. 3, Mar. 2010, doi: 10.1371/journal.pone.0009490.

[40] R. C. Edgar, “MUSCLE: Multiple sequence alignment with high accuracy and high throughput,” Nucleic Acids Res, vol. 32, no. 5, pp. 1792–1797, 2004, doi: 10.1093/nar/gkh340.

[41] T. C. Bruen, H. Philippe, and D. Bryant, “A simple and robust statistical test for detecting the presence of recombination,” Genetics, vol. 172, no. 4, pp. 2665–2681, Apr. 2006, doi: 10.1534/genetics.105.048975.

[42] I. Letunic and P. Bork, “Interactive tree of life (iTOL) v5: An online tool for phylogenetic tree display and annotation,” Nucleic Acids Res, vol. 49, no. W1, pp. W293–W296, Jul. 2021, doi: 10.1093/nar/gkab301.

[43] A. Dunai et al., “Rapid decline of bacterial drug-resistance in an antibiotic-free environment through phenotypic reversion,” 2019, doi: 10.7554/eLife.47088.001.

[44] S. Rawlinson, L. Ciric, and E. Cloutman-Green, “How to carry out microbiological sampling of healthcare environment surfaces? A review of current evidence,” Journal of Hospital Infection, vol. 103, no. 4. W.B. Saunders Ltd, pp. 363–374, Dec. 01, 2019. doi: 10.1016/j.jhin.2019.07.015.

[45] J. D. Wohrley and A. H. Bartlett, “The Role of the Environment and Colonization in Healthcare-Associated Infections,” in *Healthcare-Associated Infections in Children*, Springer International Publishing, 2019, pp. 17–36. doi: 10.1007/978-3-319-98122-2_2.

[46] W. Yin et al., “Ways to control harmful biofilms: prevention, inhibition, and eradication,” Critical Reviews in Microbiology, vol. 47, no. 1. Taylor and Francis Ltd., pp. 57–78, 2021. doi: 10.1080/1040841X.2020.1842325.

[47] M. M. Mustapha et al., “Genomic Diversity of Hospital-Acquired Infections Revealed through Prospective Whole-Genome Sequencing-Based Surveillance,” mSystems, vol. 7, no. 3, Jun. 2022, doi: 10.1128/msystems.01384-21.

[48] J. Yoon, C. H. Kim, S. Y. Yoon, C. S. Lim, and C. K. Lee, “Application of a multiplex immunochromatographic assay for rapid identification of carbapenemases in a clinical microbiology laboratory: performance and turn-around-time evaluation of NG-test Carba 5,” BMC Microbiol, vol. 21, no. 1, Dec. 2021, doi: 10.1186/s12866-021-02309-9.

[49] K. Saito et al., “Evaluation of NG-Test CARBA 5 for the detection of carbapenemase-producing Gram-negative bacilli,” J Med Microbiol, vol. 71, no. 6, 2022, doi: 10.1099/jmm.0.001557.

[50] J. Takissian, R. A. Bonnin, T. Naas, and L. Dortet, “NG-test carba 5 for rapid detection of carbapenemase-producing Enterobacterales from positive blood cultures,” Antimicrob Agents Chemother, vol. 63, no. 5, May 2019, doi: 10.1128/AAC.00011-19.

[51] A. Vasilakopoulou, P. Karakosta, S. Vourli, E. Kalogeropoulou, and S. Pournaras, “Detection of kpc, ndm and vim-producing organisms directly from rectal swabs by a multiplex lateral flow immunoassay,” Microorganisms, vol. 9, no. 5, May 2021, doi: 10.3390/microorganisms9050942.

[52] C. Shankar et al., “Identification of plasmids by PCR based replicon typing in bacteremic Klebsiella pneumoniae,” Microb Pathog, vol. 148, Nov. 2020, doi: 10.1016/j.micpath.2020.104429.

[53] W. Zhou et al., “Antimicrobial resistance and genomic characterization of Escherichia coli from pigs and chickens in Zhejiang, China,” Front Microbiol, vol. 13, Oct. 2022, doi: 10.3389/fmicb.2022.1018682.

[54] X. Cao et al., “Detection of a clinical carbapenem-resistant Citrobacter portucalensis strain and the dissemination of C. portucalensis in clinical settings,” J Glob Antimicrob Resist, vol. 27, pp. 79–81, Dec. 2021, doi: 10.1016/j.jgar.2021.04.027.

