## Supplementary materials for "Tracing the transmission of carbapenem-resistant *Enterobacterales* at the patient:ward environmental nexus"

### 1 Bioproject clinical isolate numbers

CPE = carbapenemase producing Enterobacteriales

| Patient number | Isolate number | Sample type | Species | Sequencing depth | Bioproject number | Bioproject Sample ID |
|---|---|---|---|---|---|---|
| Z1 | OXA001 | Anal CPE screen | *C. portucalensis* | 102 | PRJEB76684 | ERS20274729 |
| Z10 | OXA014 | Anal CPE screen | *E. coli* | 144 | PRJEB76684 | ERS20274730 |
| Z11 | OXA015 | Rectal CPE screen | *E. coli* | 143 | PRJEB76684 | ERS20274731 |
| Z12 | OXA016 | CPE screen | *E. coli* | 73 | PRJEB76684 | ERS20274732 |
| Z13 | OXA017 | CPE screen | *E. coli* | 146 | PRJEB76684 | ERS20274733 |
| Z14 | OXA018 | CPE screen | *E. coli* | 98 | PRJEB76684 | ERS20274734 |
| Z15 | OXA020 | Anal CPE screen | *E. hormaechei* | 148 | PRJEB76684 | ERS20274735 |
| Z15 | OXA021 | Pus (arm) | *E. hormaechei* | 148 | PRJEB76684 | ERS20274736 |
| Z15 | OXA022 | Tissue (muscle) | *E. hormaechei* | 136 | PRJEB76684 | ERS20274737 |
| Z15 | OXA023 | CPE screen | *E. hormaechei* | 103 | PRJEB76684 | ERS20274738 |
| Z16 | OXA024 | Tissue (right foot biopsy) | *E. hormaechei* | 147 | PRJEB76684 | ERS20274739 |
| Z17 | OXA025 | Anal CPE screen | *K. pneumoniae* | 50 | PRJEB76684 | ERS20274740 |
| Z18 | OXA002 | CPE screen | *K. pneumoniae* | 41 | PRJEB76684 | ERS20274741 |
| Z19 | OXA003 | CPE screen | *K. pneumoniae* | 64 | PRJEB76684 | ERS20274742 |
| Z2 | OXA005 | CPE screen | *K. pneumoniae* | 102 | PRJEB76684 | ERS20274743 |
| Z2 | OXA012 | CPE screen | *E. coli* | 94 | PRJEB76684 | ERS20274744 |
| Z20 | OXA004 | CPE screen | *K. pneumoniae* | 128 | PRJEB76684 | ERS20274745 |
| Z3 | OXA006 | Anal CPE screen | *K. pneumoniae* | 144 | PRJEB76684 | ERS20274746 |
| Z4 | OXA007 | Anal CPE screen | *K. pneumoniae* | 54 | PRJEB76684 | ERS20274747 |
| Z5 | OXA008 | Perineum CPE screen | *K. pneumoniae* | 91 | PRJEB76684 | ERS20274748 |
| Z6 | OXA009 | Anal CPE screen | *K. pneumoniae* | 105 | PRJEB76684 | ERS20274749 |
| Z6 | OXA027 | Anal CPE screen | *K. pneumoniae* | 94 | PRJEB76684 | ERS20274750 |
| Z7 | OXA010 | CPE screen | *E. coli* | 65 | PRJEB76684 | ERS20274751 |
| Z9 | OXA013 | CPE screen | *E. coli* | 101 | PRJEB76684 | ERS20274752 |
| Z9 | OXA019 | CPE screen | *E. coli* | 78 | PRJEB76684 | ERS20274753 |

#### 2 Bioproject environmental isolate numbers

WoW = workstation on wheels, HWB = hand wash basin, DWT = drain waste trap

| Isolate number | Environmental sample number | Environmental sample type | Room | Sample site | Species | Sequencing depth | Bioproject number | Bioproject sample ID |
|---|---|---|---|---|---|---|---|---|
| EI004 | 15 | Sponge swab | Sluice | Domestic Waste skip lid | *E. hormaechei* | 60 | PRJEB76684 | ERS20274754 |
| EI008 | 41 | Sponge swab | Drug Prep Room | Sink drain U bend exterior | *E. hormaechei* | 104 | PRJEB76684 | ERS20274755 |
| EI022 | 142 | Stick swab | Bay (Beds 15-18) | Bathroom toilet bowl | *C. freundii* | 141 | PRJEB76684 | ERS20274756 |
| EI026 | 158 | Stick swab | Bay (Beds 21-24) | Bathroom toilet bowl | *K. michigenensis* | 125 | PRJEB76684 | ERS20274757 |
| EI027 | 158 | Stick swab | Bay (Beds 21-24) | Bathroom toilet bowl | *C. portucalensis* | 101 | PRJEB76684 | ERS20274758 |
| EI038 | 35 | Water sample | Pantry (Staff & Patient use) | Drinks Cooler, mixed, Pre-Flush | *E. hormaechei* | 94 | PRJEB76684 | ERS20274759 |
| EI055 | 60 | Sponge swab | Workstation on wheels | WoW Keyboard and Mouse | *E. hormaechei* | 62 | PRJEB76684 | ERS20274760 |
| EI061 | 75 | Sponge swab | Bay (Beds 1-4) | Bathroom toilet floor | *C. freundii* | 126 | PRJEB76684 | ERS20274761 |
| EI071 | 74 | Stick swab | Bay (Beds 1-4) | Bathroom toilet bowl | *E. coli* | 52 | PRJEB76684 | ERS20274762 |
| EI083 | 97 | Stick swab | Bed 9 | Bathroom HWB Drain | *C. freundii* | 121 | PRJEB76684 | ERS20274763 |
| EI096 | 176 | Water sample | Staff toilet (Near Ward Office) | Staff toilet HWB DWT | *E. coli* | 270 | PRJEB76684 | ERS20274764 |
| EI097 | 177 | Sponge swab | Bed 9 | Bathroom HWB Drain | *E. coli* | 292 | PRJEB76684 | ERS20274765 |
| EI101 | 179 | Sponge swab | Pantry (Staff & Patient use) | Sink; drain waste trap; | *E. asburiae* | 83 | PRJEB76684 | ERS20274766 |
| EI106 | 178 | Water sample | Pantry (Staff & Patient use) | Sink DWT | *E. coli* | 135 | PRJEB76684 | ERS20274767 |
| EI107 | 178 | Water sample | Pantry (Staff & Patient use) | Sink DWT | *K. michigenensis* | 69 | PRJEB76684 | ERS20274768 |
| EI110 | 179 | Sponge swab | Pantry (Staff & Patient use) | Sink DWT | *K. grimontii* | 50 | PRJEB76684 | ERS20274769 |
| EI111 | 182 | Water sample | Room 33 | Office HWB DWT | *E. coli* | 62 | PRJEB76684 | ERS20274770 |
| EI113 | 182 | Water sample | Room 33 | Office HWB DWT | *C. portucalensis* | 62 | PRJEB76684 | ERS20274771 |
| EI115 | 184 | Water sample | Bay (Beds 1-4) | HWB DWT | *C. freundii* | 91 | PRJEB76684 | ERS20274772 |
| EI120 | 212 | Water sample | Bed 20 | Bathroom HWB DWT | *C. youngae* | 40 | PRJEB76684 | ERS20274773 |
| EI121 | 213 | Sponge swab | Bed 20 | HWB DWT | *C. freundii* | 76 | PRJEB76684 | ERS20274774 |
| EI122 | 212 | Water sample | Bed 20 | Bathroom HWB DWT | *E. asburiae* | 94 | PRJEB76684 | ERS20274775 |
| EI127 | 212 | Water sample | Bed 20 | HWB DWT | *C. youngae* | 68 | PRJEB76684 | ERS20274776 |
| EI129 | 182 | Water sample | Office | HWB DWT | *E. coli* | 99 | PRJEB76684 | ERS20274777 |
| EI131 | 183 | Sponge swab | Office | HWB DWT | *E. coli* | 56 | PRJEB76684 | ERS20274778 |
| EI132 | 184 | Water sample | Bay (Beds 1-4) | HWB DWT | *K. pneumoniae* | 63 | PRJEB76684 | ERS20274779 |
| EI134 | 185 | Sponge swab | Bay (Beds 1-4) | HWB DWT | *C. freundii* | 156 | PRJEB76684 | ERS20274780 |
| EI135 | 186 | Water sample | Bay (Beds 5-8) | HWB DWT | *K. pneumoniae* | 101 | PRJEB76684 | ERS20274781 |
| EI136 | 186 | Water sample | Bay (Beds 5-8) | HWB DWT | *C. freundii* | 83 | PRJEB76684 | ERS20274782 |
| EI137 | 187 | Sponge swab | Bay (Beds 5-8) | HWB DWT | *K. pneumoniae* | 98 | PRJEB76684 | ERS20274783 |

| | | | | | | | | |
|---|---|---|---|---|---|---|---|---|
| EI138 | 207 | Sponge swab | Bay (Beds 29-32) | HWB DWT | *C. youngae* | 104 | PRJEB76684 | ERS20274784 |
| EI142 | 190 | Water sample | Bed 9 | HWB DWT | *K. pneumoniae* | 52 | PRJEB76684 | ERS20274785 |
| EI145 | 194 | Water sample | Bed 10 | HWB DWT | *E. hormaechei* | 62 | PRJEB76684 | ERS20274786 |
| EI147 | 198 | Water sample | Bay (Beds 15-18) | HWB DWT | *K. michigenensis* | 78 | PRJEB76684 | ERS20274787 |
| EI148 | 199 | Sponge swab | Bay (Beds 15-18) | HWB DWT | *E. cloacae* | 52 | PRJEB76684 | ERS20274788 |
| EI153 | 203 | Sponge swab | Bed 19 | HWB DWT | *K. pneumoniae* | 41 | PRJEB76684 | ERS20274789 |
| EI154 | 203 | Sponge swab | Bed 19 | HWB DWT | *E. coli* | 137 | PRJEB76684 | ERS20274790 |
| EI155 | 206 | Water sample | Bay (Beds 29-32) | HWB DWT | *C. freundii* | 53 | PRJEB76684 | ERS20274791 |
| EI157 | 207 | Sponge swab | Bay (Beds 29-32) | HWB DWT | *C. freundii* | 96 | PRJEB76684 | ERS20274792 |
| EI160 | 208 | Water sample | Bay (Beds 29-32) | Bathroom HWB DWT | *C. freundii* | 81 | PRJEB76684 | ERS20274793 |
| EI161 | 208 | Water sample | Bay (Beds 29-32) | Bathroom HWB DWT | *K. pneumoniae* | 44 | PRJEB76684 | ERS20274794 |
| EI163 | 190 | Water sample | Bed 9 | HWB DWT | *E. roggenkampii* | 46 | PRJEB76684 | ERS20274795 |
| EI165 | 194 | Water sample | Bed 10 | HWB DWT | *C. freundii* | 71 | PRJEB76684 | ERS20274796 |
| EI166 | 195 | Sponge swab | Bed 10 | HWB DWT | *C. freundii* | 55 | PRJEB76684 | ERS20274797 |
| EI167 | 198 | Water sample | Bay (Beds 15-18) | HWB DWT | *E. coli* | 52 | PRJEB76684 | ERS20274798 |
| EI168 | 202 | Water sample | Bed 19 | HWB DWT | *E. coli* | 51 | PRJEB76684 | ERS20274799 |
| EI170 | 203 | Sponge swab | Bed 19 | HWB DWT | *C. youngae* | 78 | PRJEB76684 | ERS20274800 |
| EI171.1 | 198 | Water sample | Bay (Beds 15-18) | HWB DWT | *E. cloacae* | 65 | PRJEB76684 | ERS20274801 |
| EI171.2 | 198 | Water sample | Bay (Beds 15-18) | HWB DWT | *K. pneumoniae* | 56 | PRJEB76684 | ERS20274802 |
| EI172.1 | 200 | Water sample | Bay (Beds 15-18) | Bathroom HWB DWT | *Citrobacter sp.* | 121 | PRJEB76684 | ERS20274803 |
| EI172.2 | 200 | Water sample | Bay (Beds 15-18) | Bathroom HWB DWT | *C. youngae* | 84 | PRJEB76684 | ERS20274804 |
| EI174 | 206 | Water sample | Bay (Beds 29-32) | HWB DWT | *C. freundii* | 67 | PRJEB76684 | ERS20274805 |
| EI175 | 184 | Water sample | Bay (Beds 1-4) | HWB DWT | *P. mirabilis* | 66 | PRJEB76684 | ERS20274806 |
| EI182 | 186 | Water sample | Bay (Beds 5-8) | HWB DWT | *K. pneumoniae* | 40 | PRJEB76684 | ERS20274807 |
| EI183 | 198 | Water sample | Bay (Beds 15-18) | HWB DWT | *E. cloacae* | 46 | PRJEB76684 | ERS20274808 |
| EI184 | 202 | Water sample | Bed 19 | HWB DWT | *C. youngae* | 41 | PRJEB76684 | ERS20274809 |
| EI190 | 199 | Sponge swab | Bay (Beds 15-18) | HWB DWT | *E. coli* | 68 | PRJEB76684 | ERS20274810 |
| EI191 | 199 | Sponge swab | Bay (Beds 15-18) | HWB DWT | *K. michigenensis* | 166 | PRJEB76684 | ERS20274811 |
| EI193 | 194 | Water sample | Bed 10 | HWB DWT | *E. asburiae* | 66 | PRJEB76684 | ERS20274812 |
| EI195 | 199 | Sponge swab | Bay (Beds 15-18) | HWB DWT | *C. youngae* | 66 | PRJEB76684 | ERS20274813 |

#### 3 Bioproject metagenomic sample numbers

WoW = workstation on wheels, HWB = hand wash basin, DWT = drain waste trap. Note that these .fastq files have had all human reads removed.

| Environmental sample number | Swab type | Room | Description | Bioproject number | Bioproject sample ID |
|---|---|---|---|---|---|
| 15 | Sponge swab | Sluice | Domestic Waste skip; lid | PRJEB76684 | ERS20274814 |
| 35 | Water sample | Pantry (Staff & Patient use) | Drinks Cooler; mixed; Pre-Flush | PRJEB76684 | ERS20274815 |
| 41 | Sponge swab | Drug Prep Room | Sink drain; U bend exterior | PRJEB76684 | ERS20274816 |
| 60 | Sponge swab | Workstation on wheels | WoW; Keyboard and Mouse | PRJEB76684 | ERS20274817 |
| 71 | Stick swab | Bay (Beds 1-4) | Bathroom; HWB; Drain | PRJEB76684 | ERS20274818 |
| 74 | Stick swab | Bay (Beds 1-4) | Bathroom; toilet bowl | PRJEB76684 | ERS20274819 |
| 75 | Sponge swab | Bay (Beds 1-4) | Bathroom; toilet floor | PRJEB76684 | ERS20274820 |
| 84 | Sponge swab | Bay (Beds 5-8) | Bay Medication Cupboard; exterior surface | PRJEB76684 | ERS20274821 |
| 97 | Stick swab | Bed 9 | Bathroom; HWB; Drain | PRJEB76684 | ERS20274822 |
| 102 | Stick swab | Bed 9 | Bathroom; toilet bowl | PRJEB76684 | ERS20274823 |
| 142 | Stick swab | Bay (Beds 15-18) | Bathroom; toilet bowl | PRJEB76684 | ERS20274824 |
| 158 | Stick swab | Bay (Beds 21-24) | Bathroom; toilet bowl | PRJEB76684 | ERS20274825 |
| 176 | Water sample | Staff toilet (Near Ward Office) | Staff toilet; HWB; drain waste trap | PRJEB76684 | ERS20274826 |
| 177 | Sponge swab | Staff toilet (Near Ward Office) | Staff toilet; HWB; drain waste trap | PRJEB76684 | ERS20274827 |
| 178 | Water sample | Pantry (Staff & Patient use) | Sink; drain waste trap; | PRJEB76684 | ERS20274828 |
| 179 | Sponge swab | Pantry (Staff & Patient use) | Sink; drain waste trap; | PRJEB76684 | ERS20274829 |
| 182 | Water sample | Room 33 | Office; HWB; drain waste trap | PRJEB76684 | ERS20274830 |
| 183 | Sponge swab | Room 33 | Office; HWB; drain waste trap | PRJEB76684 | ERS20274831 |
| 184 | Water sample | Bay (Beds 1-4) | HWB; drain waste trap | PRJEB76684 | ERS20274832 |
| 185 | Sponge swab | Bay (Beds 1-4) | HWB; drain waste trap | PRJEB76684 | ERS20274833 |
| 186 | Water sample | Bay (Beds 5-8) | HWB; drain waste trap | PRJEB76684 | ERS20274834 |
| 187 | Sponge swab | Bay (Beds 5-8) | HWB; drain waste trap | PRJEB76684 | ERS20274835 |
| 190 | Water sample | Bed 9 | HWB; drain waste trap | PRJEB76684 | ERS20274836 |
| 191 | Sponge swab | Bed 9 | HWB; drain waste trap | PRJEB76684 | ERS20274837 |
| 194 | Water sample | Bed 10 | HWB; drain waste trap | PRJEB76684 | ERS20274838 |
| 195 | Sponge swab | Bed 10 | HWB; drain waste trap | PRJEB76684 | ERS20274839 |
| 198 | Water sample | Bay (Beds 15-18) | HWB; drain waste trap | PRJEB76684 | ERS20274840 |
| 199 | Sponge swab | Bay (Beds 15-18) | HWB; drain waste trap | PRJEB76684 | ERS20274841 |
| 202 | Water sample | Bed 19 | HWB; drain waste trap | PRJEB76684 | ERS20274842 |

| | | | | | |
|---|---|---|---|---|---|
| 203 | Sponge swab | Bed 19 | HWB; drain waste trap | PRJEB76684 | ERS20274843 |
| 206 | Water sample | Bay (Beds 29-32) | HWB; drain waste trap | PRJEB76684 | ERS20274844 |
| 207 | Sponge swab | Bay (Beds 29-32) | HWB; drain waste trap | PRJEB76684 | ERS20274845 |
| 208 | Water sample | Bay (Beds 29-32) | Bathroom; HWB; drain waste trap | PRJEB76684 | ERS20274846 |
| 209 | Sponge swab | Bay (Beds 29-32) | Bathroom; HWB; drain waste trap | PRJEB76684 | ERS20274847 |
| 212 | Water sample | Bed 20 | Bathroom; HWB; drain waste trap | PRJEB76684 | ERS20274848 |
| 213 | Sponge swab | Bed 20 | HWB; drain waste trap | PRJEB76684 | ERS20274849 |

### 4 Full list of $\mathit{bla}_{OXA}$ and $\mathit{bla}_{NDM}$ genes found in clinical isolates

| Clinical isolate number | Species | blaOXA-48 | blaOXA-181 | blaOXA-232 | blaOXA-244 | blaOXA-484 | blaOXA-519 | blaNDM-1 | blaNDM-2 | blaNDM-3 | blaNDM-4 | blaNDM-5 | blaNDM-6 | blaNDM-9 | blaNDM-11 |
|---|---|---|---|---|---|---|---|---|---|---|---|---|---|---|---|
| OXA001 | *C. portucalensis* | 1 | | 1 | | | | 1 | | 1 | | | | 1 | 1 |
| OXA002 | *K. pneumoniae* | | 1 | 1 | | 1 | | | | | | 1 | | | |
| OXA003 | *K. pneumoniae* | | 1 | 1 | | 1 | | | | | | 1 | | | |
| OXA004 | *K. pneumoniae* | 1 | | 1 | 1 | | 1 | 1 | 1 | 1 | | | | 1 | 1 |
| OXA005 | *K. pneumoniae* | | | 1 | | | | 1 | 1 | 1 | 1 | | | 1 | |
| OXA006 | *K. pneumoniae* | | 1 | 1 | | 1 | | | | | 1 | 1 | | | |
| OXA007 | *K. pneumoniae* | | 1 | 1 | | 1 | | | | | | 1 | | | |
| OXA008 | *K. pneumoniae* | | | | | | | | | | | 1 | | | |
| OXA009 | *K. pneumoniae* | | 1 | 1 | | 1 | | | | | | 1 | | | |
| OXA010 | *E. coli* | | 1 | | | 1 | | | | | | | | | |
| OXA012 | *E. coli* | | | | | | | 1 | | | | | | | |
| OXA013 | *E. coli* | | 1 | | | 1 | | | | | | | | | |
| OXA014 | *E. coli* | | 1 | | | 1 | | | | | | | | | |
| OXA015 | *E. coli* | | 1 | | | 1 | | | | | | | | | |
| OXA016 | *E. coli* | | 1 | | | | | | | | | | | | |
| OXA017 | *E. coli* | | 1 | | | 1 | | | | | | | | | |
| OXA018 | *E. coli* | | 1 | | | 1 | | | | | | | | | |
| OXA019 | *E. coli* | | 1 | | | 1 | | | | | | | | | |
| OXA020 | *E. hormaechei* | | | | | | | 1 | | | | | | | |
| OXA021 | *E. hormaechei* | | | | | | | 1 | | | | | | 1 | |
| OXA022 | *E. hormaechei* | | | | | | | 1 | 1 | 1 | | | | 1 | 1 |
| OXA023 | *E. hormaechei* | | | | | | | 1 | | | | | | | |
| OXA024 | *E. hormaechei* | | | | | | | 1 | | | | | 1 | 1 | |
| OXA026 | *K. pneumoniae* | | 1 | 1 | | 1 | | | | | | 1 | | | |
| OXA027 | *K. pneumoniae* | | 1 | 1 | | 1 | | | | | 1 | 1 | | | |
| | Number | 2 | 15 | 10 | 1 | 14 | 1 | 9 | 3 | 4 | 3 | 8 | 1 | 6 | 3 |
| | Percentage | 8% | 58% | 38% | 4% | 54% | 4% | 35% | 12% | 15% | 12% | 31% | 4% | 23% | 12% |

#### 5 Full list of *bla*$_{OXA}$ and *bla*$_{NDM}$ genes found in environmental isolates

| Isolate number | Species | blaOXA-48 | blaOXA-181 | blaOXA-232 | blaOXA-244 | blaOXA-484 | blaOXA-519 | blaNDM-1 | blaNDM-2 | blaNDM-3 | blaNDM-4 | blaNDM-5 | blaNDM-6 | blaNDM-9 | blaNDM-11 |
|---|---|---|---|---|---|---|---|---|---|---|---|---|---|---|---|
| 22 | *C. freundii* | 1 | 1 | 1 | | | 1 | | | | | | | | |
| 61 | *C. freundii* | | | | | | | | | | | | | | |
| 83 | *C. freundii* | | | | | | | | | | | | | | |
| 115 | *C. freundii* | | | | | | | | | | | | | | |
| 121 | *C. freundii* | | | | | | | | | | | | | | |
| 134 | *C. freundii* | | | | | | | | | | | | | | |
| 136 | *C. freundii* | | | | | | | | | | | | | | |
| 155 | *C. freundii* | 1 | | | 1 | | 1 | | | | | | | | |
| 157 | *C. freundii* | 1 | 1 | | | | 1 | | | | | | | | |
| 160 | *C. freundii* | 1 | 1 | 1 | 1 | | 1 | | | | | | | | |
| 165 | *C. freundii* | | | | | | | | | | | | | | |
| 166 | *C. freundii* | | | | | | | | | | | | | | |
| 174 | *C. freundii* | 1 | 1 | 1 | | | | | | | | | | | |
| 27 | *C. portucalensis* | 1 | | 1 | 1 | | | | | | | | | | |
| 113 | *C. portucalensis* | 1 | | 1 | | | | | | | | | | | |
| 120 | *C. youngae* | | | | | | | | | | | | | | |
| 127 | *C. youngae* | | | | | | | | | | | | | | |
| 138 | *C. youngae* | | | | | | | | | | | | | | |
| 170 | *C. youngae* | | | | | | | | | | | | | | |
| 172.2 | *C. youngae* | | | | | | | | | | | | | | |
| 184 | *C. youngae* | 1 | | | | | 1 | | | | | | | | |
| 195 | *C. youngae* | | | | | | | | | | | | | | |
| 172.1 | *Citrobacter sp.* | | | | | | | | | | | | | | |
| 101 | *E. asburiae* | | | | | | | | | | | | | | |
| 122 | *E. asburiae* | | | | | | | | | | | | | | |
| 193 | *E. asburiae* | | | | | | | | | | | | | | |
| 148 | *E. cloacae* | | | | | | | | | | | | | | |
| 171.1 | *E. cloacae* | | | | | | | | | | | | | | |
| 183 | *E. cloacae* | | | | | | | | | | | | | | |

| | | | | | | | | | | | | | | | |
|---|---|---|---|---|---|---|---|---|---|---|---|---|---|---|---|
| 71 | *E. coli* | | | | | | | | | | | | | | |
| 96 | *E. coli* | | | | | | | | | | | | | | |
| 97 | *E. coli* | | | | | | | | | | | | | | |
| 106 | *E. coli* | | | | | | | | | | | | | | |
| 111 | *E. coli* | | | | | | | | | | | | | | |
| 129 | *E. coli* | | | | | | | | | | | | | | |
| 131 | *E. coli* | | | | | | | | | | | | | | |
| 154 | *E. coli* | | | | | | | | | | | | | | |
| 167 | *E. coli* | | | | | | | | | | | | | | |
| 168 | *E. coli* | | | | | | | | | | | | | | |
| 190 | *E. coli* | | | | | | | | | | | | | | |
| 4 | *E. hormaechei* | | | | | | | | | | | | | | |
| 8 | *E. hormaechei* | | | | | | | | | | | | | | |
| 38 | *E. hormaechei* | | | | | | | | | | | | | | |
| 55 | *E. hormaechei* | | | | | | | | | | | | | | |
| 145 | *E. hormaechei* | | | | | | | | | | | | | | |
| 163 | *E. roggenkampii* | | | | | | | | | | | | | | |
| 110 | *K. grimontii* | | | | | | | | | | | | | | |
| 26 | *K. michigenensis* | 1 | | | | | 1 | 1 | | | | | | | |
| 107 | *K. michigenensis* | | | | | | | | | | | | | | |
| 147 | *K. michigenensis* | | | | | | | | | | | | | | |
| 191 | *K. michigenensis* | | | | | | | | | | | | | | |
| 132 | *K. pneumoniae* | | | | | | | | | | | | | | |
| 135 | *K. pneumoniae* | | | | | | | | | | | | | | |
| 137 | *K. pneumoniae* | | | | | | | | | | | | | | |
| 142 | *K. pneumoniae* | | | | | | | | | | | | | | |
| 153 | *K. pneumoniae* | 1 | | 1 | | | | | | | | | | | |
| 161 | *K. pneumoniae* | | | | | | | | | | | | | | |
| 171.2 | *K. pneumoniae* | 1 | | | | | | | | | | | | | |
| 182 | *K. pneumoniae* | | | | | | | | | | | | | | |
| 175 | *P. mirabilis* | | | | | | | | | | | | | | |
| | Total | 11 | 4 | 6 | 3 | 0 | 6 | 1 | 0 | 0 | 0 | 0 | 0 | 0 | 0 |
| | Percentage | 18% | 7% | 10% | 5% | 0% | 10% | 2% | 0% | 0% | 0% | 0% | 0% | 0% | 0% |

### 6 Plasmids found in environmental isolates

| | *C. farmeri* | *C. freundii* | *Citrobacter sp.* | *C. youngae* | *E. asburiae* | *E. cloacae* | *E. cloacae complex* | *E. hormaechei* | *Enterobacter sp.* | *E. coli* | *K. oxytoca* | *K. pneumoniae* | **Total** |
|---|---|---|---|---|---|---|---|---|---|---|---|---|---|
| Col(IMGS31) | 0 (0.0%) | 2 (15.4) | 0 (0.0%) | 0 (0.0%) | 0 (0.0%) | 1 (20.0%) | 1 (33.3%) | 0 (0.0%) | 1 (100%) | 3 (25.0%) | 3 (75.0%) | 0 (0.0%) | 11 (18.6%) |
| Col(IRGK) | 1 (100%) | 5 (38.5%) | 2 (66.7%) | 1 (100%) | 2 (100%) | 1 (20.0%) | 2 66.7%) | 0 (0.0%) | 1 (100%) | 10 (83.3%) | 2 (50.0%) | 8 (61.5%) | 35 (59.3%) |
| Col(pHAD28) | 0 (0.0%) | 1 (7.7%) | 2 (66.7%) | 1 (100%) | 0 (0.0%) | 2 (40.0%) | 2 66.7%) | 0 (0.0%) | 0 (0.0%) | 4 (33.3%) | 1 (25.0%) | 0 (0.0%) | 13 (22.0%) |
| Col156 | 1 (100%) | 5 (38.5%) | 2 (66.7%) | 0 (0.0%) | 0 (0.0%) | 2 (40.0%) | 2 66.7%) | 0 (0.0%) | 0 (0.0%) | 6 (50.0%) | 2 (50.0%) | 4 (30.8%) | 24 (40.7%) |
| Col440I | 0 (0.0%) | 0 (0.0%) | 0 (0.0%) | 0 (0.0%) | 0 (0.0%) | 0 (0.0%) | 0 (0.0%) | 0 (0.0%) | 0 (0.0%) | 0 (0.0%) | 2 (50.0%) | 0 (0.0%) | 2 (3.4%) |
| ColRNAI | 0 (0.0%) | 1 (7.7%) | 0 (0.0%) | 0 (0.0%) | 0 (0.0%) | 0 (0.0%) | 0 (0.0%) | 0 (0.0%) | 0 (0.0%) | 0 (0.0%) | 1 (25.0%) | 0 (0.0%) | 2 (3.4%) |
| IncC | 0 (0.0%) | 2 (15.4) | 0 (0.0%) | 1 (100%) | 0 (0.0%) | 0 (0.0%) | 0 (0.0%) | 0 (0.0%) | 0 (0.0%) | 0 (0.0%) | 1 (25.0%) | 0 (0.0%) | 4 (6.8%) |
| IncFIA(HI1) | 1 (100%) | 0 (0.0%) | 0 (0.0%) | 0 (0.0%) | 0 (0.0%) | 1 (20.0%) | 0 (0.0%) | 0 (0.0%) | 0 (0.0%) | 2 (16.7%) | 1 (25.0%) | 0 (0.0%) | 5 (8,5%) |
| IncFIA(pBK30683) | 0 (0.0%) | 0 (0.0%) | 0 (0.0%) | 0 (0.0%) | 0 (0.0%) | 0 (0.0%) | 1 (33.3%) | 0 (0.0%) | 0 (0.0%) | 0 (0.0%) | 0 (0.0%) | 1 (7.7%) | 2 (3.4%) |
| IncFIB(AP001918) | 0 (0.0%) | 0 (0.0%) | 0 (0.0%) | 0 (0.0%) | 0 (0.0%) | 0 (0.0%) | 0 (0.0%) | 0 (0.0%) | 0 (0.0%) | 1 (8.3%) | 0 (0.0%) | 0 (0.0%) | 1 (1.7%) |
| IncFIB(K) | 0 (0.0%) | 1 (7.7%) | 1 (33.3%) | 0 (0.0%) | 0 (0.0%) | 3 (60.0%) | 0 (0.0%) | 1 (100%) | 0 (0.0%) | 6 (50.0%) | 2 (50.0%) | 13 (100%) | 27 (45.8%) |
| IncFIB(K)(pCAV1099-114) | 0 (0.0%) | 0 (0.0%) | 0 (0.0%) | 0 (0.0%) | 0 (0.0%) | 0 (0.0%) | 0 (0.0%) | 0 (0.0%) | 0 (0.0%) | 0 (0.0%) | 1 (25.0%) | 0 (0.0%) | 1 (1.7%) |
| IncFIB(pECLA) | 0 (0.0%) | 1 (7.7%) | 0 (0.0%) | 0 (0.0%) | 0 (0.0%) | 1 (20.0%) | 1 (33.3%) | 0 (0.0%) | 1 (100%) | 0 (0.0%) | 1 (25.0%) | 0 (0.0%) | 7 (11.9%) |
| IncFIB(pNDM-Mar) | 0 (0.0%) | 0 (0.0%) | 0 (0.0%) | 0 (0.0%) | 0 (0.0%) | 0 (0.0%) | 0 (0.0%) | 0 (0.0%) | 0 (0.0%) | 0 (0.0%) | 2 (50.0%) | 0 (0.0%) | 2 (3.4%) |
| IncFIB(pQil) | 0 (0.0%) | 0 (0.0%) | 0 (0.0%) | 0 (0.0%) | 0 (0.0%) | 0 (0.0%) | 0 (0.0%) | 0 (0.0%) | 0 (0.0%) | 0 (0.0%) | 1 (25.0%) | 1 (7.7%) | 2 (3.4%) |
| IncFII | 0 (0.0%) | 1 (7.7%) | 0 (0.0%) | 0 (0.0%) | 0 (0.0%) | 1 (20.0%) | 0 (0.0%) | 0 (0.0%) | 0 (0.0%) | 5 (41.7%) | 0 (0.0%) | 0 (0.0%) | 7 (11.9%) |
| IncFII(29) | 0 (0.0%) | 0 (0.0%) | 0 (0.0%) | 0 (0.0%) | 0 (0.0%) | 0 (0.0%) | 0 (0.0%) | 0 (0.0%) | 0 (0.0%) | 1 (8.3%) | 0 (0.0%) | 0 (0.0%) | 1 (1.7%) |
| IncFII(Cf) | 0 (0.0%) | 1 (7.7%) | 0 (0.0%) | 0 (0.0%) | 0 (0.0%) | 0 (0.0%) | 0 (0.0%) | 0 (0.0%) | 0 (0.0%) | 0 (0.0%) | 0 (0.0%) | 0 (0.0%) | 1 (1.7%) |

| | | | | | | | | | | | | | |
|---|---|---|---|---|---|---|---|---|---|---|---|---|---|
| IncFII(K) | 0 (0.0%) | 0 (0.0%) | 1 (33.3%) | 0 (0.0%) | 0 (0.0%) | 2 (40.0%) | 0 (0.0%) | 0 (0.0%) | 0 (0.0%) | 1 (8.3%) | 3 (75.0%) | 10 (76.9%) | 17 (28.8%) |
| Inc(pBK30683) | 0 (0.0%) | 0 (0.0%) | 0 (0.0%) | 0 (0.0%) | 0 (0.0%) | 0 (0.0%) | 0 (0.0%) | 0 (0.0%) | 0 (0.0%) | 0 (0.0%) | 0 (0.0%) | 1 (7.7%) | 1 (1.7%) |
| IncFII(pCRY) | 0 (0.0%) | 1 (7.7%) | 0 (0.0%) | 0 (0.0%) | 0 (0.0%) | 0 (0.0%) | 0 (0.0%) | 0 (0.0%) | 0 (0.0%) | 0 (0.0%) | 0 (0.0%) | 0 (0.0%) | 1 (1.7%) |
| IncFII(pECLA) | 0 (0.0%) | 0 (0.0%) | 0 (0.0%) | 0 (0.0%) | 1 (50.0%) | 1 (20.0%) | 0 (0.0%) | 0 (0.0%) | 1 (100%) | 0 (0.0%) | 1 (25.0%) | 0 (0.0%) | 4 (6.8%) |
| IncFII(SARC14) | 0 (0.0%) | 0 (0.0%) | 0 (0.0%) | 0 (0.0%) | 0 (0.0%) | 0 (0.0%) | 0 (0.0%) | 0 (0.0%) | 0 (0.0%) | 0 (0.0%) | 1 (25.0%) | 0 (0.0%) | 1 (1.7%) |
| IncFII(Yp) | 0 (0.0%) | 1 (7.7%) | 0 (0.0%) | 0 (0.0%) | 0 (0.0%) | 0 (0.0%) | 0 (0.0%) | 1 (100%) | 0 (0.0%) | 0 (0.0%) | 2 (50.0%) | 0 (0.0%) | 4 (6.8%) |
| IncHI1A | 0 (0.0%) | 0 (0.0%) | 0 (0.0%) | 0 (0.0%) | 0 (0.0%) | 0 (0.0%) | 0 (0.0%) | 0 (0.0%) | 0 (0.0%) | 3 (25.0%) | 0 (0.0%) | 0 (0.0%) | 3 (5.1%) |
| IncHI1A(NDM-CIT) | 0 (0.0%) | 2 (15.4) | 0 (0.0%) | 0 (0.0%) | 0 (0.0%) | 0 (0.0%) | 0 (0.0%) | 0 (0.0%) | 0 (0.0%) | 0 (0.0%) | 0 (0.0%) | 0 (0.0%) | 2 (3.4%) |
| IncHI1B(pNDM-CIT) | 0 (0.0%) | 2 (15.4) | 0 (0.0%) | 0 (0.0%) | 0 (0.0%) | 0 (0.0%) | 0 (0.0%) | 0 (0.0%) | 0 (0.0%) | 0 (0.0%) | 0 (0.0%) | 0 (0.0%) | 2 (3.4%) |
| IncHI1B(R27) | 0 (0.0%) | 0 (0.0%) | 0 (0.0%) | 0 (0.0%) | 0 (0.0%) | 0 (0.0%) | 0 (0.0%) | 0 (0.0%) | 0 (0.0%) | 3 (25.0%) | 1 (25.0%) | 0 (0.0%) | 4 (6.8%) |
| IncHI2 | 0 (0.0%) | 0 (0.0%) | 0 (0.0%) | 0 (0.0%) | 1 (50.0%) | 1 (20.0%) | 1 (33.3%) | 0 (0.0%) | 0 (0.0%) | 0 (0.0%) | 0 (0.0%) | 0 (0.0%) | 3 (5.1%) |
| IncHI2A | 0 (0.0%) | 0 (0.0%) | 0 (0.0%) | 0 (0.0%) | 1 (50.0%) | 1 (20.0%) | 1 (33.3%) | 0 (0.0%) | 0 (0.0%) | 0 (0.0%) | 0 (0.0%) | 0 (0.0%) | 3 (5.1%) |
| IncI1-I(alpha) | 0 (0.0%) | 0 (0.0%) | 0 (0.0%) | 0 (0.0%) | 0 (0.0%) | 0 (0.0%) | 0 (0.0%) | 0 (0.0%) | 0 (0.0%) | 1 (8.3%) | 0 (0.0%) | 0 (0.0%) | 1 (1.7%) |
| IncL | 0 (0.0%) | 6 (46.2%) | 0 (0.0%) | 0 (0.0%) | 0 (0.0%) | 0 (0.0%) | 0 (0.0%) | 0 (0.0%) | 0 (0.0%) | 0 (0.0%) | 1 (25.0%) | 5 (38.5%) | 12 (20.3%) |
| IncM1 | 0 (0.0%) | 0 (0.0%) | 0 (0.0%) | 1 (100%) | 1 (50.0%) | 1 (20.0%) | 1 (33.3%) | 0 (0.0%) | 0 (0.0%) | 1 (8.3%) | 1 (25.0%) | 0 (0.0%) | 6 (10.2%) |
| IncN | 0 (0.0%) | 1 (7.7%) | 0 (0.0%) | 0 (0.0%) | 0 (0.0%) | 0 (0.0%) | 0 (0.0%) | 0 (0.0%) | 0 (0.0%) | 2 (16.7%) | 0 (0.0%) | 0 (0.0%) | 3 (5.1%) |
| IncQ2 | 0 (0.0%) | 0 (0.0%) | 1 (33.3%) | 0 (0.0%) | 0 (0.0%) | 0 (0.0%) | 0 (0.0%) | 0 (0.0%) | 0 (0.0%) | 0 (0.0%) | 0 (0.0%) | 0 (0.0%) | 1 (1.7%) |
| IncP6 | 0 (0.0%) | 0 (0.0%) | 0 (0.0%) | 0 (0.0%) | 0 (0.0%) | 0 (0.0%) | 0 (0.0%) | 0 (0.0%) | 0 (0.0%) | 3 (25.0%) | 0 (0.0%) | 0 (0.0%) | 3 (5.1%) |
| IncX1 | 0 (0.0%) | 5 (38.5%) | 0 (0.0%) | 0 (0.0%) | 0 (0.0%) | 0 (0.0%) | 0 (0.0%) | 0 (0.0%) | 0 (0.0%) | 0 (0.0%) | 0 (0.0%) | 0 (0.0%) | 5 (8,5%) |
| IncX5 | 0 (0.0%) | 0 (0.0%) | 1 (33.3%) | 0 (0.0%) | 0 (0.0%) | 0 (0.0%) | 0 (0.0%) | 0 (0.0%) | 0 (0.0%) | 0 (0.0%) | 0 (0.0%) | 0 (0.0%) | 1 (1.7%) |

| | | | | | | | | | | | | | |
|---|---|---|---|---|---|---|---|---|---|---|---|---|---|
| IncY | 0 (0.0%) | 0 (0.0%) | 0 (0.0%) | 0 (0.0%) | 0 (0.0%) | 0 (0.0%) | 0 (0.0%) | 0 (0.0%) | 0 (0.0%) | 4 (33.3%) | 0 (0.0%) | 0 (0.0%) | 4 (6.8%) |
| pENTAS02 | 0 (0.0%) | 0 (0.0%) | 0 (0.0%) | 0 (0.0%) | 0 (0.0%) | 2 (40.0%) | 0 (0.0%) | 0 (0.0%) | 0 (0.0%) | 0 (0.0%) | 0 (0.0%) | 0 (0.0%) | 2 (3.4%) |
| pKPC-CAV | 0 (0.0%) | 1 (7.7%) | 0 (0.0%) | 0 (0.0%) | 0 (0.0%) | 0 (0.0%) | 0 (0.0%) | 0 (0.0%) | 0 (0.0%) | 0 (0.0%) | 0 (0.0%) | 0 (0.0%) | 1 (1.7%) |
| pKPC-CAV1321 | 1 (100%) | 8 (62.5%) | 2 (66.7%) | 1 (100%) | 1 (50.0%) | 0 (0.0%) | 0 (0.0%) | 0 (0.0%) | 0 (0.0%) | 0 (0.0%) | 1 (25.0%) | 2 (15.4%) | 16 (21.1%) |
| repB(R1701) | 0 (0.0%) | 1 (7.7%) | 1 (33.3%) | 0 (0.0%) | 1 (50.0%) | 3 (60.0%) | 0 (0.0%) | 0 (0.0%) | 0 (0.0%) | 0 (0.0%) | 3 (75.0%) | 3 (23.1%) | 12 (10.2%) |

### 7 Environmental metagenomic samples – Enterobacterial plasmids

Table split in half to accommodate all plasmids. WoW = workstation on wheels, HWB = hand wash basin, DWT = drain waste trap

| Environmental sample site | Room | Description | Total plasmids per sample | Col(IMGS31) | ColKP3 | Col(IRGK) | Col156 | ColRNAI | IncA | IncC | IncFIA | IncFIA(HI1) | IncFIA(HI1)(pAR0022) | IncFIA(pBK30683) | IncFIB(AP001918) | IncFIB(K) | IncFIB(K)(pCAV1099-114) | IncFIB(pECLA) | IncFIB(pHCM2) | IncFIB(pQil) | IncFII | IncFII(Cf) | Inc(pBK30683) | IncFII(pECLA) |
|---|---|---|---|---|---|---|---|---|---|---|---|---|---|---|---|---|---|---|---|---|---|---|---|---|
| 15 | Sluice | Domestic Waste skip; lid | 0 | | | | | | | | | | | | | | | | | | | | | |
| 35 | Pantry | Drinks Cooler; Pre-Flush | 0 | | | | | | | | | | | | | | | | | | | | | |
| 41 | Drug Prep Room | Sink drain; U bend exterior | 1 | | | | | | | | | | | | | | | | | | | | | |
| 60 | WoW | WoW; Keyboard and Mouse | 0 | | | | | | | | | | | | | | | | | | | | | |
| 71 | Bay (Beds 1-4) | Bathroom; HWB; Drain | 0 | | | | | | | | | | | | | | | | | | | | | |
| 74 | Bay (Beds 1-4) | Bathroom; toilet bowl | 5 | | | | 1 | | | | 1 | | | | 1 | | | | | | | | | |
| 75 | Bay (Beds 1-4) | Bathroom; toilet floor | 0 | | | | | | | | | | | | | | | | | | | | | |
| 84 | Bay (Beds 5-8) | Bay Medication Cupboard | 0 | | | | | | | | | | | | | | | | | | | | | |
| 97 | Bed 9 | Bathroom; HWB; Drain | 0 | | | | | | | | | | | | | | | | | | | | | |
| 102 | Bed 9 | Bathroom; toilet bowl | 0 | | | | | | | | | | | | | | | | | | | | | |
| 142 | Bay (Beds 15-18) | Bathroom; toilet bowl | 1 | | | | | | | | | | | | | | | | | | | | | |
| 158 | Bay (Beds 21-24) | Bathroom; toilet bowl | 5 | | | | | | | 1 | | | | | | 1 | | | | | | 1 | | |
| 176 | Staff toilet | Staff toilet; HWB; DWT | 2 | | | 1 | 1 | | | | | | | | | | | | | | | | | |
| 177 | Staff toilet | Staff toilet; HWB; DWT | 7 | | | | 1 | | | | | 1 | | | | 1 | 1 | | | | 1 | | | |
| 178 | Pantry | Sink; DWT | 0 | | | | | | | | | | | | | | | | | | | | | |
| 179 | Pantry | Sink; DWT | 0 | | | | | | | | | | | | | | | | | | | | | |
| 182 | Room 33 | Office; HWB; DWT | 2 | | | | | | | | | | | | | | | | | | | | | |
| 183 | Room 33 | Office; HWB; DWT | 3 | | | | | | | | | | | | | | | | | | | | | |
| 184 | Bay (Beds 1-4) | HWB; DWT | 3 | | | | | | | | | | | | | 1 | | | | | | | | |
| 185 | Bay (Beds 1-4) | HWB; DWT | 2 | | | | | | | | | | | | | | | | | | | | | |
| 186 | Bay (Beds 5-8) | HWB; DWT | 5 | 1 | | | | | | | | | | | | 1 | | | | | | | | |
| 187 | Bay (Beds 5-8) | HWB; DWT | 10 | 1 | | | | 1 | | | | | 1 | | | | | | | | | | | 1 |
| 190 | Bed 9 | HWB; DWT | 5 | 1 | | | | | | | | | | | | 1 | | | | | | | | |

| | | | | | | | | | | | | | | | | | | | | | | | | |
|---|---|---|---|---|---|---|---|---|---|---|---|---|---|---|---|---|---|---|---|---|---|---|---|---|
| 191 | Bed 9 | HWB; DWT | 4 | 1 | | | | | | | | | | | | | | | | | | | | |
| 194 | Bed 10 | HWB; DWT | 1 | | | | | | | | | | | | | | | | | | | | | |
| 195 | Bed 10 | HWB; DWT | 14 | 1 | | 1 | | | | 1 | | | 1 | | | 1 | | 1 | 1 | | 1 | 1 | | |
| 198 | Bay (Beds 15-18) | HWB; DWT | 6 | | | 1 | | | | | | | | | | 1 | | | | | 1 | | 1 | |
| 199 | Bay (Beds 15-18) | HWB; DWT | 12 | 1 | | 1 | | 1 | 1 | 1 | | | 1 | | | 1 | | | 1 | | 1 | | | |
| 202 | Bed 19 | HWB; DWT | 7 | | 1 | | | | | 1 | | | | 1 | | | | 1 | | 1 | | | | |
| 203 | Bed 19 | HWB; DWT | 6 | | 1 | | | | | 1 | | | | 1 | | | | | | | | | | |
| 206 | Bay (Beds 29-32) | HWB; DWT | 7 | | | | | | | | | | | | | 1 | | 1 | | | | | | 1 |
| 207 | Bay (Beds 29-32) | HWB; DWT | 8 | | | | | | | | | 1 | | | | 1 | | 1 | | | | 1 | | 1 |
| 208 | Bay (Beds 29-32) | Bathroom; HWB; DWT | 0 | | | | | | | | | | | | | | | | | | | | | |
| 209 | Bay (Beds 29-32) | Bathroom; HWB; DWT | 0 | | | | | | | | | | | | | | | | | | | | | |
| 212 | Bed 20 | Bathroom; HWB; DWT | 3 | | | | | | | | | | | | | | | 1 | | | | | | |
| 213 | Bed 20 | HWB; DWT | 3 | | | | | | 1 | 1 | | | | | | | | | | | | | | |
| | | | Total | 6 | 2 | 4 | 3 | 2 | 2 | 6 | 1 | 2 | 3 | 2 | 1 | 10 | 1 | 5 | 2 | 1 | 4 | 3 | 1 | 3 |
| | | | Percentage | 8% | 3% | 5% | 4% | 3% | 3% | 8% | 1% | 3% | 4% | 3% | 1% | 13% | 1% | 7% | 3% | 1% | 5% | 4% | 1% | 4% |

| Environmental sample site | Room | Description | Total plasmids per sample | IncFII(pKPX1) | IncFII(pAR0022) | IncFII(Yp) | IncHI1A | IncHI1A(NDM-CIT) | IncHI1B(pNDM-CIT) | IncHI2 | IncHI2A | IncI1-I(alpha) | IncM1 | IncN3 | IncQ2 | IncP1 | IncR | IncX1 | IncX5 | IncX6 | IncY | pKPC-CAV1321 | repB(R1701) |
|---|---|---|---|---|---|---|---|---|---|---|---|---|---|---|---|---|---|---|---|---|---|---|---|
| 15 | Sluice | Domestic Waste skip; lid | 0 | | | | | | | | | | | | | | | | | | | | |
| 35 | Pantry | Drinks Cooler; Pre-Flush | 0 | | | | | | | | | | | | | | | | | | | | |
| 41 | Drug Prep Room | Sink drain; U bend exterior | 1 | | | | | | | | | | | | | 1 | | | | | | | |
| 60 | WoW | WoW; Keyboard and Mouse | 0 | | | | | | | | | | | | | | | | | | | | |
| 71 | Bay (Beds 1-4) | Bathroom; HWB; Drain | 0 | | | | | | | | | | | | | | | | | | | | |
| 74 | Bay (Beds 1-4) | Bathroom; toilet bowl | 5 | | | | | | | | | 1 | | | | | | | | | 1 | | |
| 75 | Bay (Beds 1-4) | Bathroom; toilet floor | 0 | | | | | | | | | | | | | | | | | | | | |

| | | | | | | | | | | | | | | | | | | | | | | | |
|---|---|---|---|---|---|---|---|---|---|---|---|---|---|---|---|---|---|---|---|---|---|---|---|
| 84 | Bay (Beds 5-8) | Bay Medication Cupboard | 0 | | | | | | | | | | | | | | | | | | | | |
| 97 | Bed 9 | Bathroom; HWB; Drain | 0 | | | | | | | | | | | | | | | | | | | | |
| 102 | Bed 9 | Bathroom; toilet bowl | 0 | | | | | | | | | | | | | | | | | | | | |
| 142 | Bay (Beds 15-18) | Bathroom; toilet bowl | 1 | | | | | | | | | | | | 1 | | | | | | | | |
| 158 | Bay (Beds 21-24) | Bathroom; toilet bowl | 5 | | | | | | | | | | | | | | | 1 | | | | 1 | |
| 176 | Staff toilet | Staff toilet; HWB; DWT | 2 | | | | | | | | | | | | | | | | | | | | |
| 177 | Staff toilet | Staff toilet; HWB; DWT | 7 | | | | 1 | | | | | | | | | | | | | | | | 1 |
| 178 | Pantry | Sink; DWT | 0 | | | | | | | | | | | | | | | | | | | | |
| 179 | Pantry | Sink; DWT | 0 | | | | | | | | | | | | | | | | | | | | |
| 182 | Room 33 | Office; HWB; DWT | 2 | | | | | 1 | | | | | | | | | 1 | | | | | | |
| 183 | Room 33 | Office; HWB; DWT | 3 | | | | | 1 | 1 | | | | | | | | 1 | | | | | | |
| 184 | Bay (Beds 1-4) | HWB; DWT | 3 | | | | | 1 | 1 | | | | | | | | | | | | | | |
| 185 | Bay (Beds 1-4) | HWB; DWT | 2 | | | | | 1 | 1 | | | | | | | | | | | | | | |
| 186 | Bay (Beds 5-8) | HWB; DWT | 5 | | | | | | | | | | | | | | | | 1 | | | 1 | 1 |
| 187 | Bay (Beds 5-8) | HWB; DWT | 10 | | 1 | | | | | | | | 1 | | | | | | 1 | 1 | | 1 | 1 |
| 190 | Bed 9 | HWB; DWT | 5 | | | | | | | 1 | 1 | | | | | | 1 | | | | | | |
| 191 | Bed 9 | HWB; DWT | 4 | | | | | | | 1 | 1 | | | | | | 1 | | | | | | |
| 194 | Bed 10 | HWB; DWT | 1 | | | | | | | | | | 1 | | | | | | | | | | |
| 195 | Bed 10 | HWB; DWT | 14 | | | | | | | 1 | 1 | | 1 | | | | | | | | | 1 | 1 |
| 198 | Bay (Beds 15-18) | HWB; DWT | 6 | | | | | | | | | | | | | | | | 1 | | | | 1 |
| 199 | Bay (Beds 15-18) | HWB; DWT | 12 | | | | | | | | | | | | | | | | 1 | | | 1 | 1 |
| 202 | Bed 19 | HWB; DWT | 7 | | | | | | | | | | | | | | | | | | | 1 | 1 |
| 203 | Bed 19 | HWB; DWT | 6 | | | 1 | | | | | | | | 1 | | | | | | | | 1 | |
| 206 | Bay (Beds 29-32) | HWB; DWT | 7 | 1 | | 1 | | | | | | | | | | | | | | | | 1 | 1 |
| 207 | Bay (Beds 29-32) | HWB; DWT | 8 | | | 1 | | | | | | | | | | | | | | | | 1 | 1 |
| 208 | Bay (Beds 29-32) | Bathroom; HWB; DWT | 0 | | | | | | | | | | | | | | | | | | | | |
| 209 | Bay (Beds 29-32) | Bathroom; HWB; DWT | 0 | | | | | | | | | | | | | | | | | | | | |
| 212 | Bed 20 | Bathroom; HWB; DWT | 3 | | | | | | | | | | | | | | | | | | | 1 | 1 |
| 213 | Bed 20 | HWB; DWT | 3 | | | | | | | | | | | | | | | | | | | 1 | |
| | | | Total | 1 | 1 | 3 | 1 | 4 | 3 | 3 | 3 | 1 | 3 | 1 | 1 | 1 | 4 | 1 | 4 | 1 | 1 | 11 | 10 |
| | | | Percentage | 1% | 1% | 4% | 1% | 5% | 4% | 4% | 4% | 1% | 4% | 1% | 1% | 1% | 5% | 1% | 5% | 1% | 1% | 15% | 13% |
